# Treatment outcome is associated with pre-treatment connectome measures across psychiatric disorders - evidence for connectomic reserve?

**DOI:** 10.1101/2025.02.23.25322741

**Authors:** Chris Vriend, Sophie M.D.D. Fitzsimmons, Inga Aarts, Aniek Broekhuizen, Ysbrand D. van der Werf, Linda Douw, Henny A. D. Visser, Kathleen Thomaes, Odile A. van den Heuvel

## Abstract

**Background:** Predicting treatment efficacy in psychiatric disorders remains challenging, despite the availability of effective interventions. Previous studies suggest a link between brain network topology and treatment efficacy in individual disorders, but cross-disorder investigations are lacking.

**Methods:** We analyzed pre-treatment MRI data from 177 individuals (113 females) with either obsessive-compulsive disorder (OCD) or post-traumatic stress disorder with comorbid personality disorders (PTSD) that received different non-pharmacological treatments. Using diffusion and resting-state MRI, we constructed structural, functional, and multilayer connectomes and calculated network measures at the global and mesoscale for network integration (global efficiency, eccentricity), segregation (modularity) and their balance (small-worldness). We assessed the relationship between these pre-treatment network measures and treatment improvement using mixed-model and Bayesian analyses and compared them to healthy controls. We also investigated associations between response and treatment-induced changes in network measures.

**Results:** Across disorders and treatments, psychiatric cases showed a 41.6±29.6% symptom improvement (62% response rate) after treatment and pre-treatment differences in functional and multilayer network topology compared to healthy controls. Symptom improvement was associated with pre-treatment functional (P=0.04) and structural small-worldness (P=0.01), and multilayer eccentricity (P=0.01), while responders had higher functional modularity (P=0.02). Results were robust across trials and treatments, when adjusting for medication status and showed high credibility in Bayesian analyses. Network change associations with treatment response were only modest.

**Conclusions:** Pre-treatment connectome characteristics are related to treatment response, regardless of treatment and psychiatric disorder, and suggest that individual differences in intrinsic features of the human connectome underlie amenability to treatment.

## INTRODUCTION

Deficits in cognitive control – the ability to regulate one’s thoughts, emotions and behavior – underlie many of the cognitive and psychological symptoms in individuals with a psychiatric disorder.(1,2) For example, anxiety provoking thoughts (obsessions) and ritualistic behaviors (compulsions) in patients with obsessive-compulsive disorder (OCD), or traumatic flashbacks in post-traumatic stress disorder (PTSD) are associated with cognitive control deficits. Across psychiatric disorders, these cognitive control deficits are accompanied by disturbed communication within the brain ‘connectome’: the arrangement of structural and functional connections between brain areas that form a complex network.(3) According to graph theory, the normal architecture of the connectome is governed by two fundamental principles: segregation (processing in specialized subsystems, e.g. visual system) and integration (i.e. exchange between subsystems).(3) A key component of this architecture is also a modular network structure with sparse cross-modular long range connections.(4) It has previously been suggested that the development and severity of brain-related disorders is associated with an imbalance between segregation and integration (5). This view is supported by studies that have suggested common connectome dysfunction across brain-related disorders, both in psychiatry,(4,6,7) and neurology.(8,9) Nevertheless, few head-to-head comparisons between brain-related disorders have been conducted. Some studies have investigated cross-disorder differences in local brain anatomy.(9) Others showed structural dysconnectivity of brain areas involved in cognitive control that are also vital for network integration,(8) or common functional patterns across psychiatric disorders with only subtle differences with healthy controls.(6)

Treatments, such as psychotherapy and neurostimulation, can enhance cognitive control and have been shown to normalize connectome dysfunction.(10–12) Unfortunately, however, not everyone (fully) benefits from these treatments,(13) with significant risk of chronicity. Predicting who will respond to treatment remains, however, challenging. Some studies have been able to predict the outcome of cognitive behavioral therapy,(14–16) and neurostimulation,(17,18) but none of these predictors are currently used in the clinic, mainly due to replication failures.(19,20)

Motivated by the previous findings on shared deficits across psychiatric disorders in both cognitive control and the organizational structure of the connectome, this study will investigate whether cross-disorder connectome characteristics are associated with treatment outcome of non-pharmacological, cognitive control enhancing treatments. Previous studies have already identified modularity as a potential predictor for treatment efficacy in single treatments and disorders,(21) but to our knowledge this has not yet been investigated across disorders and treatments in a single study. Identification of network markers that predict treatment outcome across psychiatric disorders not only fits with the dimensional approach of Research Domain Criteria (RdoC),(22) and allows leveraging of larger, better powered and diverser datasets, it can also enhance our understanding of commonalities in treatment resistance and help tailor mechanism-based treatments by focusing more on individual variation in network topology along a continuum rather than on symptom-based diagnostic labels. We associated pre-treatment network measures with treatment response by combining data from four clinical trials with pre- and post-treatment structural and functional MRI in two psychiatric disorders (OCD and PTSD) and nine different treatments (see methods section) involving different forms of psychotherapy (some in combination with neurostimulation). We hypothesized (see also our pre-registration: <u>osf.io/p7bcn</u>) that efficacy of treatment was associated with pre-treatment network measures of segregation and integration, regardless of the psychiatric disorder or treatment. We additionally investigated cross-disorder associations between treatment-induced network changes and treatment response, hypothesizing that successful treatment would be associated with changes in network integration and segregation. Given the novelty of this research topic we did not specify hypotheses about the directionality or the importance of either modality.

## METHODS AND MATERIALS

### Participants and interventions

This project included participants from several completed clinical trials: ARRIBA (NCT03929081), TIPICCO (NCT03667807), and PROSPER (NCT03833531 / NCT03833453). Details are provided in the supplements and in the clinical trial papers (23–26). Briefly, ARRIBA involved OCD participants randomized to receive either cognitive behavioral therapy (CBT) with exposure and response prevention (ERP) or inference-based CBT (I-CBT). TIPICCO participants with OCD received ERP combined with repetitive transcranial magnetic stimulation (rTMS). Both trials measured OCD symptom severity with the Yale-Brown Obsessive-Compulsive Scale (YBOCS) before and after treatment. The PROSPER study comprised two trials for individuals with PTSD and a comorbid personality disorder: PROSPER-B and PROSPER-C. PROSPER-B participants received eye movement desensitization and reprocessing (EMDR) alone or with dialectical behavioral therapy (DBT). PROSPER-C participants received imagery rescripting alone or with schema therapy. Both trials used the clinician-administered PTSD interview for DSM-5 (CAPS-5) to assess PTSD severity pre- and post-treatment. All participants received active treatment and provided informed consent. All trials were approved by the VU Medical Center Medical Ethical Committee. Inclusion/exclusion criteria are in the supplements. Only ARRIBA, TIPICCO, and a subset of PROSPER participants underwent MRI, and those with baseline diffusion MRI, resting-state functional MRI, and pre/post-treatment clinical measures were included in the analysis. Non-compliant individuals with post-treatment assessments were included as an intention-to-treat sample. A matched healthy control group, free of psychopathology, medication and neurological disorders, that were scanned on the same MRI scanner (see below) was selected from the ARRIBA, TIPICCO, PROSPER, and additional studies, COGTIPS,(27) and the global OCD study.(28)

### Image acquisition and preprocessing

All participants were scanned on a GE Discovery MR750 3T scanner (General Electric, Milwaukee, U.S) using a 32-channel head coil with harmonized sequences, including a 3D T1-weighted MRI, resting-state fMRI (rsfMRI) and multi-shell diffusion MRI (dMRI). Acquisition parameters and preprocessing steps are provided in the supplements. FMRI scans with a mean FD>0.5 mm were excluded. DMRI scans were visually inspected for residual (motion) artifacts and if necessary excluded.

### Connectome construction and network measures

We used the Schaefer 300P7N atlas and 14 individually segmented subcortical areas (FreeSurfer 7.3.2) to extract time series from the preprocessed, denoised functional images. Following recommendations for consistent network reconstruction (29,30) we applied Pearson correlations, absolutized negative correlations, and used Orthogonal Minimal Spanning Tree (OMST) thresholding to create a 314×314 weighted functional connectome. The same parcellation and OMST thresholding algorithm were used to construct a weighted structural connectome from the dMRI tractogram. To build a multilayer connectome, we calculated the minimum spanning tree (MST) of both structural and functional connectomes and created Interlayer links (with weight 1) that connect the same nodes across structural and functional layers. The use of MST prevents differences in link density and connection strength between layers and participants.(31)

We calculated several widely used global network measures from the structural, functional and multilayer connectomes that capture the integrative and segregative capacity of connectomes. Global efficiency, average participation coefficient (PC), modularity and small-worldness were calculated for the structural and functional connectomes. Since these measures lack a suitable multilayer counterpart, we calculated the average eccentricity and eigenvector centrality instead. Definitions and details are provided in the supplements.

On the mesoscale, we investigated structural and functional connectivity strength between eight ‘systems’, seven Yeo cortical networks (32) – Default mode (DMN), Frontoparietal (FPN), Dorsal attention (DAN), Ventral Attention (VAN), Somatomotor (SMN), Limbic and Visual Network – and 14 subcortical regions. We additionally calculated the average PC of the nodes in each of the systems for the structural and functional connectomes and the average eccentricity and eigenvector centrality for the multilayer connectome (as PC is not suitable for multilayer connectomes). More details are provided in the supplements.

### Statistical analysis

Analyses were pre-registered with the Open Science Framework (osf.io/p7bcn). Because the trials used different scales as outcome measures, we defined treatment outcome across disorders as the percentage improvement from pre-to-post-treatment. We additionally defined treatment responders and non-responders based on the established cut-off score for OCD (Y-BOCS: ≥35% improvement,(33) and ≥ 1 pooled SD improvement on the CAPS-5 as no uniform response definition for PTSD treatment exists.(24)

We performed univariate mixed model analyses with pre-treatment global network measures (single or multilayer) as the dependent variable and percentage change or responder/non-responder as the independent variable, with and without covariates for age and sex. To capture consistent and universal markers of treatment response we only adjusted for clinical trial or treatment in separate sensitivity analyses using random intercepts. These null hypothesis significance testing (NHST) models were evaluated at p < 0.05, and as pre-registered, the global measures were not corrected for multiple comparisons.

At the mesoscale, we used Bayesian multilevel modeling to include information across all systems into one statistical model. Treatment outcome was associated with structural and functional connectivity between the eight systems using Matrix-based Bayesian analyses (34) and with averaged PC measures (single layers) and averaged eccentricity and eigenvector centrality measures (multilayer) of the eight systems using Region-Based Analysis Program through Bayesian Multilevel Modeling (RBA).(35). The advantages of these Bayesian multilevel approaches for neuroimaging data, including more transparent reporting and simultaneous integration of related measures from the same brain, are detailed in the supplements. Bayesian models produce posterior distributions, with the area under the curve to the right or left of the zero-effect line representing the effect’s credibility, expressed as the positive posterior probability (P+). The distance of the posterior mode from the zero-effect line indicates the effect’s magnitude. While inferences are based on the full posterior distribution, we only report effects with P+ <0.10 or >0.90 in the main text and classified effect credibility as *moderate* (P+ between [1-]0.05 and [1-]0.10), *strong* (P+ between [1-]0.01 and [1-]0.05), and very strong (P+ <0.01 or >0.99). We also performed exploratory network-based statistics (NBS) on the full 314×314 structural or functional connectomes to identify clusters of edges that formed an interconnected subnetwork that are associated with treatment outcome at a T-threshold of 3.1 (default).

Additionally, using similar statistical models, we compared the psychiatric samples at baseline with the pooled healthy control sample to investigate the pre-treatment starting point relative to healthy controls. We used the MatchIt R package to select age and sex matched controls from a pool of N=158.

To determine how single-layer and multilayer topology of the network change with successful treatment we used models that deviated from the pre-registration plan. These models and the reasons for the deviations are explained in the supplements.

Lastly, leave-one-trial out and leave-one-treatment out (collectively called leave-one-sample-out [LOSO]) validation were performed to determine the robustness of the findings. For the global topology measures we calculated the harmonic mean p-value across folds which can handle dependencies between p-values and is less stringent than a Bonferroni correction.(36) For the Bayesian analyses we report the median and range in P+ values across folds.

### Code availability

Preprocessing and analyses code is available at: github.com/chrisvriend/core-multi

## RESULTS

### Demographics, clinical information

Of the 200 individuals that completed the interventions, 23 had to be excluded because they did not consent to the re-use of their data, leaving data from 177 for the current project. Forty-three participants did not return for the post-treatment MRI (in part because of COVID regulations), leaving 134 for the pre-to-post treatment analyses (supplementary table 1). Some additional functional or diffusion MRI scans had to be excluded because of missing or low quality data (see flowchart in supplementary figure 1). The prediction sample included 172 individuals with diffusion MRI, 167 with functional MRI and 162 individuals with both. The demographic and clinical details of the prediction sample are presented in Table 1, showing on average a 41.6 (±29.6) percent treatment-induced symptom improvement across trials and a 61.6% response rate. Before treatment, 87 were on psychotropic medication. Medicated and non-medicated individuals did not differ in percentage improvement (t_(155)_ = 0.26, P=0.80) or response rate (X_(1)_= 0.024, P = 0.99). There were no differences in frame-wise displacement during fMRI between the treatment groups (F_(8,166)_ = 0.69, P=0.70). MatchIt selected 143 healthy controls that were matched on age (Welch’s t_(283.84)_=0.77, P=0.44) and sex (χ_(1)_=3.3, P=0.07) and they showed no differences in frame-wise displacement (F_(1,312)_ = 0.08 P=0.78).

**Table 1.** Demographic and clinical characteristics.

| Sample | Treatment | N | Age (yrs) | T0 sev.# | T1 sev.# | % improvement | Sex (F/M) | Responder (no/yes) | Med (no/yes) | FD fMRI |
| --- | --- | --- | --- | --- | --- | --- | --- | --- | --- | --- |
| **Total** |  | 177 | 36.7 ± 11.7 |  |  | 41.6 ± 29.6 | 113/64 | 68/109 | 88/87* |  |
| ARRIBA | CBT-ERP | 28 | 30.4 ± 9.1 | 24 ± 3.7 | 12.2 ± 7.2 | 48.1 ± 33.6 | 15/13 | 10/18 | 15/13 | 0.12 ± 0.05 |
|  | I-CBT | 35 | 34 ± 10.5 | 24.2 ± 4.4 | 16.3 ± 7.1 | 32.9 ± 25.3 | 19/16 | 18/17 | 19/16 | 0.14 ± 0.06 |
| TIPICCO | DLPFC | 17 | 41.5 ± 12.2 | 28.5 ± 5.7 | 16.8 ± 8.1 | 41.9 ± 22.8 | 11/6 | 6/11 | 5/12 | 0.15 ± 0.09 |
|  | preSMA | 20 | 32.5 ± 14 | 29 ± 3.5 | 19.6 ± 7.9 | 33.2 ± 25 | 11/9 | 12/8 | 7/13 | 0.13 ± 0.06 |
|  | vertex | 18 | 40.1 ± 11.6 | 26.6 ± 4 | 15.6 ± 4.8 | 40.5 ± 19.5 | 12/6 | 6/12 | 9/9 | 0.12 ± 0.05 |
| PROSPER-B | EMDR | 14 | 36.1 ± 10.9 | 47.8 ± 10.2 | 30.8 ± 15.6 | 37.6 ± 28.2 | 13/1 | 4/10 | 8/6 | 0.12 ± 0.04 |
|  | EMDR+DBT | 11 | 37.7 ± 7.2 | 41.7 ± 11 | 23.9 ± 14.4 | 41.9 ± 35.1 | 9/2 | 4/7 | 6/4 | 0.11 ± 0.04 |
| PROSPER-C | IR | 16 | 45.4 ± 11.4 | 37.8 ± 11.3 | 19 ± 18.7 | 51.2 ± 43.3 | 10/6 | 4/12 | 8/7 | 0.15 ± 0.12 |
|  | IR+SFT | 18 | 40.3 ± 10.9 | 45.3 ± 9.9 | 22.1 ± 16.6 | 53.2 ± 29.5 | 13/5 | 4/14 | 11/7 | 0.14 ± 0.06 |
| Healthy controls | - | 143 | 37.8 ± 13.4 | - | - | - | 76/67 | - | - | 0.13 ± 0.06 |

Scores are presented as mean ± standard deviation, unless otherwise indicated. # score represents the severity on the Yale-Brown Obsessive-compulsive Scale (Y-BOCS) for the ARRIBA and TIPICCO trials, and the clinician-administered PTSD scale for DSM-5 (CAPS-5) for the PROSPER trials. Medication is operationalized as using selective serotonin reuptake inhibitors (SSRI's), serotonin-norepinephrine reuptake inhibitors (SNRI), tricyclic antidepressants (TCAs), antipsychotics or mood stabilizers at the time of the T0 MRI scan. \* data was missing for two cases. Abbreviations: CBT-ERP = cognitive behavioral therapy (CBT) with exposure and response prevention (ERP) therapy, IBA = inference based approach, DLPCF = repetitive Transcranial Magnetic Stimulation (rTMS) to the dorsolateral prefrontal cortex in combination with ERP, preSMA = rTMS to the pre-supplementary motor area in combination with ERP, vertex = rTMS to the vertex in combination with ERP, EMDR = eye-movement desensitization and reprocessing, EMDR+DBT = EMDR and dialectical behavioral therapy, IR = imagery rescripting therapy, IR+SFT = imagery rescripting and schema focused therapy.

### Pre-treatment case-control differences

Results are reported in the supplements, showing differences in global and system topology of the functional and multilayer, but not structural connectome (Figure 1, supplementary Tables 2-3, supplementary Figures 2-3). These differences suggest that psychiatric cases have suboptimal integrative and segregative properties of the functional connectome and lower multilayer integrative capacity.

**Figure 1.**
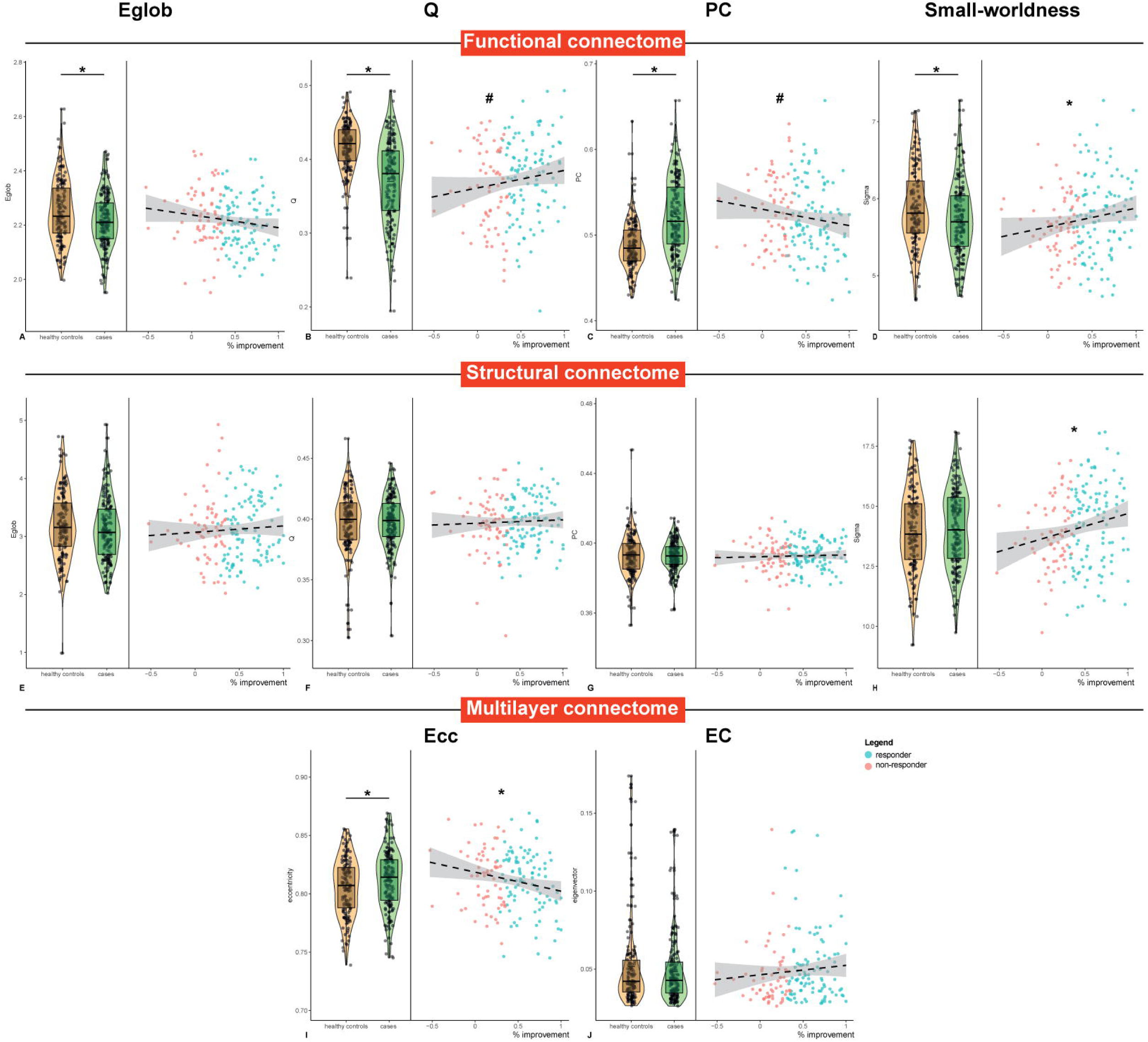
Case-control differences and associations with treatment outcome of pre-treatment global network measure. Violin plots show the distribution of the functional (a-d), structural (e-h) and multilayer (i-j) network measures. There were significant between-group differences in all functional network measures and the average multilayer eccentricity. Scatterplots show the associations between pre-treatment network measures and percentage symptom improvement, with positive values representing improvement after treatment. individual dots are colored according to whether someone was a responder (green) or non-responder (red). Percentage symptom improvement was significantly associated with functional (d) and structural (h) small-worldness and multilayer average eccentricity (i). Responders and non-responders also showed significant differences in functional modularity (b) and average participation coefficient (c). * significant association between percentage symptom improvement and the pre-treatment network measures. # significant difference in pre-treatment network measure between treatment responders and non-responders. abbreviations: Eglob = global efficiency, Q = modularity, PC = average participation coefficient, Ecc = average eccentricity, EC = average eigenvector centrality.

### Treatment outcome and pre-treatment network measures

Percentage symptom improvement was positively associated with pre-treatment small-worldness of the functional (B[SE]=24.4[12.6], P=0.04) and structural connectome (B[SE]=105.4[41.4], P=0.01) and negatively with multilayer eccentricity (B[SE]=−1.6[0.7], P=0.01). Compared with non-responders, responders showed a higher modularity (B[SE]=1.8[0.9], P=0.02) and lower average PC (B[SE]=−1.3[0.7], P=0.03) of the functional connectome (Supplementary Table 4). Results were robust against LOSO validation (Supplementary Table 4) and sensitivity analyses adjusting for trial or treatment showed similar results (Supplementary Table 5). Other global measures were not associated with treatment response (supplementary Table 6).

On the mesoscale, there was moderate evidence for a negative association between percentage improvement and functional, but not structural connectivity of the DMN with the FPN, DAN and limbic network (P+ = 0.08-0.09; Supplementary Figure 4). The association with DMN-FPN connectivity was most robust. Compared with non-responders, responders showed moderately credible evidence for lower functional connectivity between the DMN and FPN (P+ = 0.06) and DAN (P+ = 0.06) and lower structural DMN-FPN, DAN-VAN, DAN-FPN and VAN-SMN (P+ = 0.05-0.09) connectivity. There was strong to very strong credibility for associations of percentage improvement and response status with lower PC across all functional systems and particularly for the DMN, FPN, limbic and visual networks (all P+ < 0.1; Figure 2). There was only credible evidence for higher PC of the VAN (P+ = 0.94) and subcortical areas (P+ = 1) in responders. Multilayer eccentricity showed very strongly credible evidence for a negative association with percentage improvement (all P+ <0.05) but not response status across all systems (Figure 3), while there was moderately credible evidence for higher multilayer eigenvector centrality in responders in the FPN (P+ =0.91), DAN (P+ =0.93), VAN (P+=0.92) and limbic network (P+−0.91). These results were generally robust to LOSO validation (Figures 2-3 and supplementary Figure 5-6). Exploratory NBS did not produce an interconnected subcluster associated with treatment efficacy.

**Figure 2.**
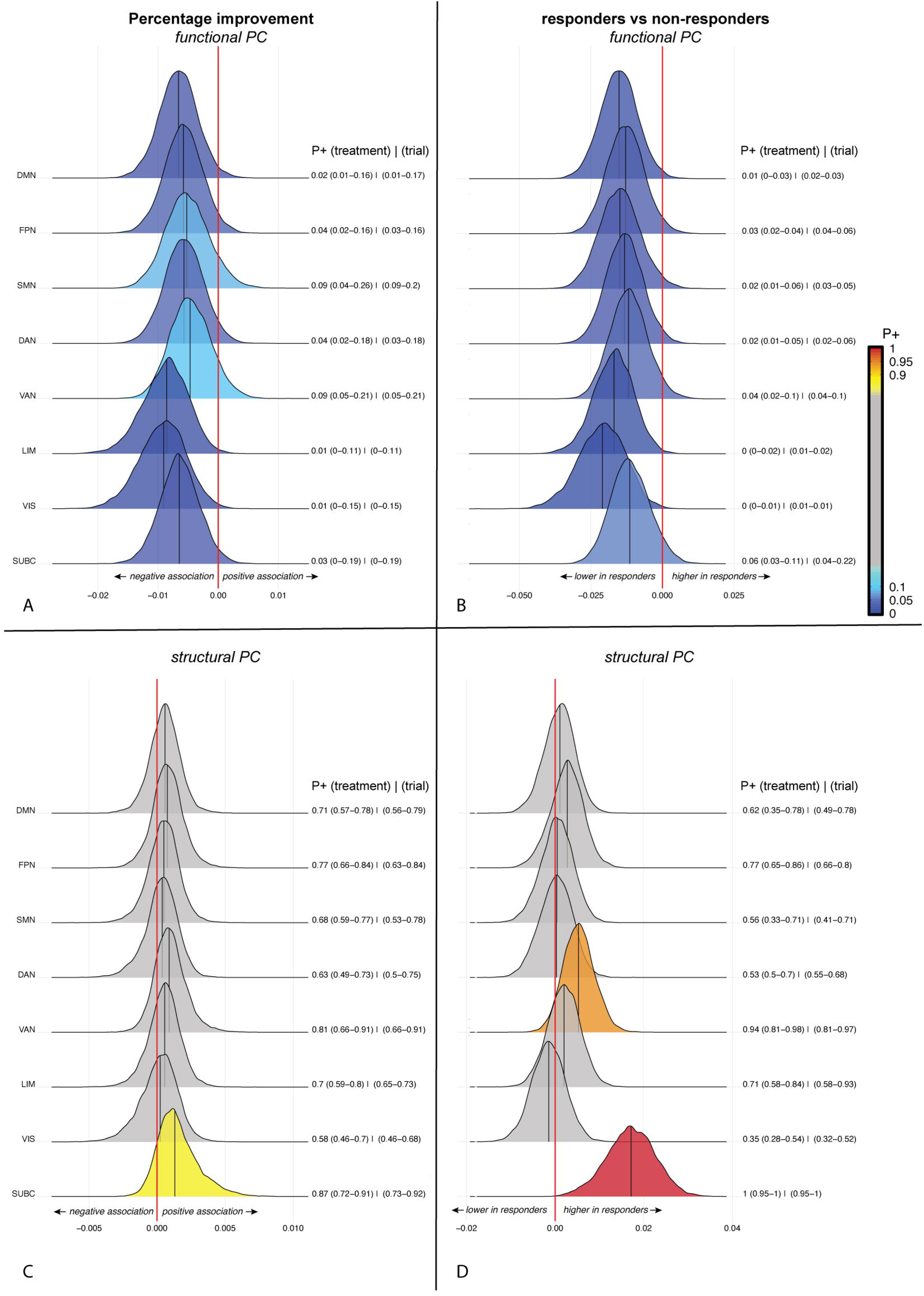
Bayesian posterior distribution plots of the association between structural and functional participation coefficient of eight brain systems and percentage change (a,b) or response status (c,d). The posterior distribution communicates the credibility of an effect. Positive posterior probabilities (*P+*) are shown next to each distribution and color coded. The values between brackets indicate the range of P+ values across leave-one-treatment-out, or leave-one-trial-out folds to show the robustness of the results. *P+* values ≥0.90 indicate moderate to very high credibility for a positive effect, P+ ≤0.10 indicate moderate to very high credibility for a negative effect. The meaning of the direction of effects are shown next to the red zero-effect line. There was credible evidence for (a) negative associations between functional participation coefficient and percentage symptom improvement, and (b) lower functional participation coefficient in responders compared with non-responders across all eight systems. With the exception of a higher structural participation coefficient of the ventral attention network and subcortical structures in responders compared with non-responders (d), there was no credible evidence for associations between treatment outcome and structural participation coefficient. abbreviations: PC = participation coefficient, DMN = default mode network, FPN = frontoparietal network, SMN = somatomotor network, DAN = dorsal attention network, VAN = ventral attention network, LIM = limbic network, VIS = visual network. SUBC = subcortical structures.

**Figure 3.**
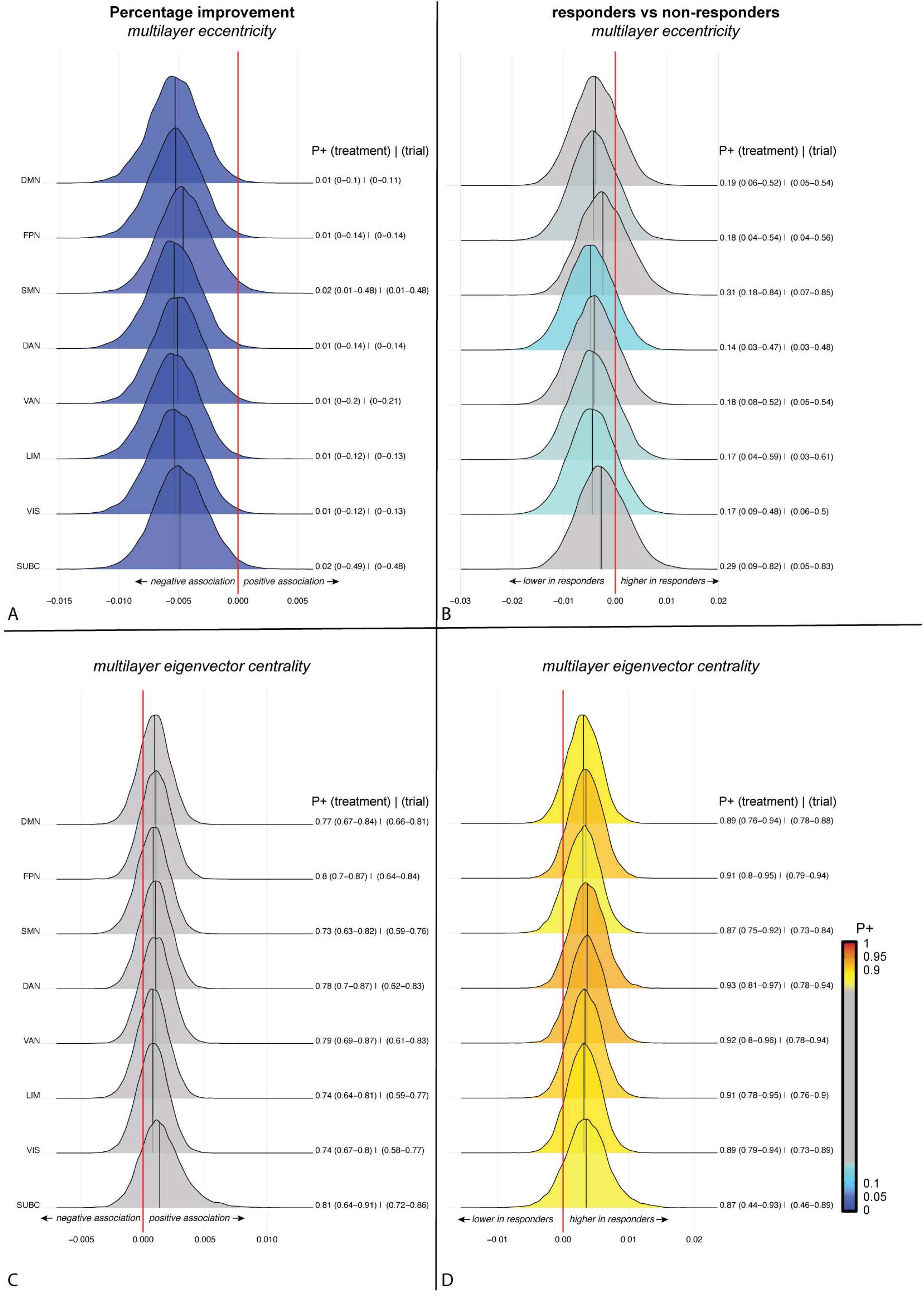
Bayesian posterior distribution plots of the associations between multilayer eigenvector centrality and eccentricity of eight brain systems and percentage change (a,b) or response status (c,d). See the legend of figure 2 for an explanation of the posterior distribution plots. There was credible evidence for (a) negative associations between multilayer eccentricity and percentage symptom improvement across all eight systems, but (b) no credible evidence for differences between responders and non-responders. There was also no credible evidence for an association between (c) symptom improvement and multilayer eigenvector centrality. (d) multilayer eigenvector centrality was higher in responders compared with non-responders in all eight networks but only the dorsal and ventral attention network as well as frontoparietal and limbic network showed a P+ > 0.90. abbreviations: DMN = default mode network, FPN = frontoparietal network, SMN = somatomotor network, DAN = dorsal attention network, VAN = ventral attention network, LIM = limbic network, VIS = visual network. SUBC = subcortical structures.

### Pre-to-post treatment changes

On the global level there were no significant associations between the change in single layer or multilayer network measures and treatment response, except that responders and non-responders differed in the pre-to-post-treatment change in small-worldness of the structural connectome (B[SE]=−61.2[25.9], P=0.02) and multilayer eigenvector centrality (B[SE]=−1.2[0.6], P=0.047; see Supplementary Table 7). Non-responders showed an increase in small-worldness and eigenvector centrality, while responders showed a decrease in small-worldness but no change in eigenvector centrality. Adjusting for trial or treatment did not affect these results and there were no changes in other global measures (Supplementary Table 8-9).

Results on the mesoscale are reported in the supplements (Supplementary Figures 7-11). NBS showed no interconnected subcluster that changed with treatment-induced symptom improvement or differed between responders and non-responders.

### Post-hoc analyses

To account for possible medication effects we added medication status as an additional covariate to the analyses associating the pre-treatment global topological measures and treatment response. Results remained, except for the small-worldness of the functional connectome which no longer showed a significant association (see supplementary Table 10).

We performed Bayesian analyses with LOSO validation on the pre-treatment global topological measures to corroborate our NHST findings (supplementary Figures 12-13). These analyses showed highly credible and robust evidence for the reported associations with treatment efficacy. In addition, it showed high credibility for associations between percentage improvement and functional global efficiency (P+=0.03), average PC (P+ =0.03), modularity (P+ = 0.96) and multilayer eigenvector centrality (P+=0.90), and similar results for responder versus non-responders.

Lastly, we repeated the analyses in the two OCD trials with five different treatments. Results are reported in the supplements (Supplementary Table 11).

## DISCUSSION

The aim of this study was to establish common connectome markers that are associated with successful treatment outcome across two psychiatric disorders (OCD and PTSD) encompassing nine different treatments from four clinical trials. Our results showed that treatment response was associated with several global measures of segregation and integration, primarily of the functional connectome, and an integration measure (eccentricity) of the structure-function multilayer. These global results were largely mirrored on the mesoscale and using complementary statistical approaches, i.e. NHST and Bayesian (multilevel) modeling, showing strong credible evidence for these associations. The results were overall robust against leave-one-trial and leave-one-treatment out validation and medication status.

Individuals with the largest imbalance between integration and segregation (expressed as a lower small-worldness) of both the functional and structural connectomes benefited the least from treatment, as did individuals with lower functional modularity (i.e. lower modularity, higher average participation coefficient) and lower integrative capacity of the structure-function multilayer (eccentricity). Together these results suggest that individuals with brain connectomes that deviate more from a balanced and ‘healthy’ connectome are the least amenable to cognitive control enhancing treatments, regardless of the disorder and treatment (Figure 4). This coincides with previous studies that identified modularity as a potential cross-disorder imaging marker for treatment response.(21,37–39)

**Figure 4.**
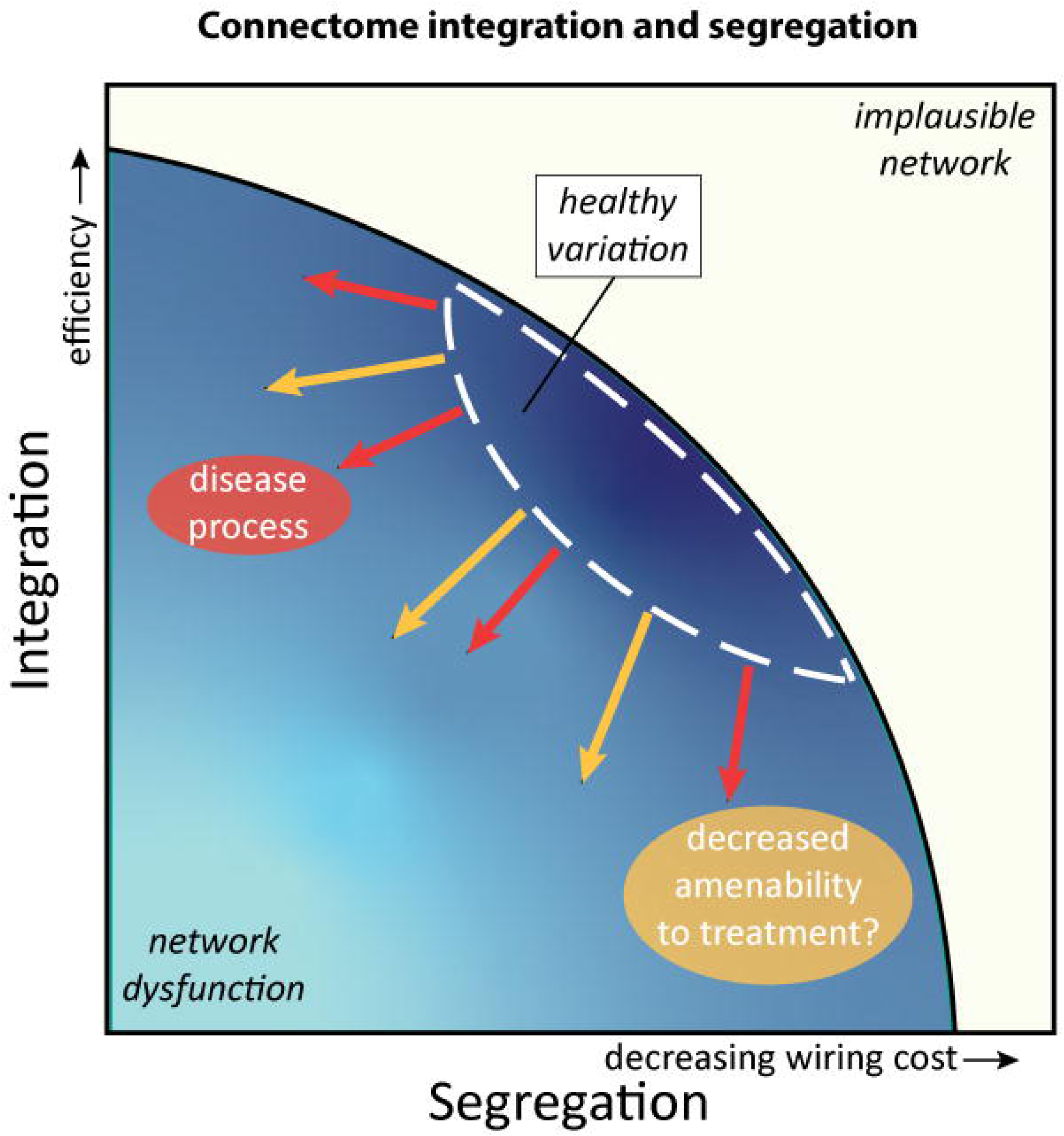
Schematic representation of the association between network topology and amenability to treatment. The architecture of the connectome is governed by the fundamental principles that under normal circumstances are in balance: segregation, i.e. tight local connections between brain areas that form a specialized subsystem, and integration, i.e. exchange between subsystems with long range connections. Deviation from the healthy variation in balance between these forces is associated with network dysfunction and the emergence of brain-related disorders as reviewed by (5). The results of the current study suggest that (further) deviation from the norm is also associated with decreased amenability to non-pharmacological treatment for psychiatric disorders.

Individual differences in the brain’s capacity to resist pathological changes – regardless of what these are – has previously been conceptualized as a cognitive reserve,(40) of which the neurobiological underpinnings are yet to be fully elucidated. Likewise, we postulate that the brain’s capacity to *benefit* from treatment can be conceptualized as a *Connectomic Reserve*: an intrinsic feature of someone’s brain network, regardless of the disorder, of which the interindividual variation determines amenability to treatment. Our current results suggest that a normal modular network structure and healthy balance between network segregation and integration is associated with greater benefits from treatment, but more research is needed to determine whether these topological measures can predict treatment outcome on the individual level.

We saw relatively few associations with treatment-induced changes in the connectivity and topology of the connectome, most of which were sensitive to LOSO validation. Both responders and non-responders showed an increase in multilayer eigenvector centrality after treatment, but this increase was higher in the non-responders. This was contrary to what we expected. Non-responders additionally showed larger decreases in structural connectivity and an increase in small-worldness after treatment compared with responders. An increase in average eigenvector centrality is indicative of higher integration and efficiency of the entire network. However, neither single layer global efficiency nor multilayer eccentricity, which are more direct measures of the efficiency of the network, showed such an effect. Instead we speculate that these findings in combination with the decrease in system-to-system structural connectivity strength and increase in small-worldness may point towards a maladaptive shift in the hierarchical organization of the network. This maladaptive shift represents more local clustering between influential brain regions but at the cost of cross-modular long range connections. The fact that on the system level we saw a relatively low robustness against LOSO validation, may stem from the smaller sample with pre-and-post-treatment MRI data but may alternatively suggest that the different treatments impact the connectome differently and increase variability.

Our findings further support previous reports showing common connectome dysfunction across psychiatric disorders (4,6,7), by showing on average a less favorable functional and multilayer network topology and lower functional connectivity in psychiatric cases compared with healthy controls. Nevertheless, there was quite some variation, not only between individuals but also between trials and treatments and some findings were no longer significant when adjusting for trial or treatment (although the findings were robust against LOSO validation).

This study has a number of strengths. First, we used harmonized state-of-the-art MRI acquisitions with stringent quality control and a uniform processing pipeline. We additionally investigated different brain network modalities at multiple scales and used two complementary statistical approaches (NHST and Bayesian modeling), LOSO validation and two definitions for treatment response (percentage change, response status) to allow investigation of the robustness of the findings. Although there were some differences in the level of statistical significance or credibility of evidence between the percentage improvement and responder/non-responder models, with the exception of the multilayer eccentricity, they all pointed in the same direction. We currently do not have an explanation for the contrasting findings for multilayer eccentricity but this may stem from a difference in variance.

Some other limitations also need to be kept in mind when interpreting these findings. The current analyses were performed on the group level in two psychiatric disorders and it remains to be determined whether the findings extend to other psychiatric (and neurological) disorders, other (non-pharmacological) treatments and whether cross-disorder connectome measures can predict treatment outcome on the individual level. Let this manuscript therefore also be an invitation to all readers to collaborate and further test the existence of a universal Connectomic Reserve. The use of imaging data acquired on the same scanner with harmonized sequences and processing pipelines also prevented us from investigating robustness against differences in scanners and processing pipelines. Although adjusting for sex (and age) did not influence the results, there was a relatively higher predominance of females in this sample which may have influenced the results.

In conclusion, we showed for the first time that treatment outcome is related to transdiagnostic pre-treatment network topology across two psychiatric disorders and nine different non-pharmacological treatments. Specifically, we showed that individual variation in intrinsic features of the human connectome related to functional network modularity, integration/segregation balance and integration across the structure-function multilayer, underlies amenability to treatment. These results support a more dimensional approach towards precision medicine for psychiatric disorders with potential for network measures as cross-disorder biological markers for treatment response.

## Data Availability

All data produced in the present study are available upon reasonable request to the authors
Preprocessing and analyses code is available at: github.com/chrisvriend/core-multi

## Acknowledgments

The current project was executed without funding. The TIPICCO trial was funded by the Netherlands Organization for Health Research Talent Programme (Vidi Grant No. 91717306 [to OAvdH]. The ARRIBA trial was supported by a grant from ZonMW (Grant No. 636310004 [to HADV]). The PROSPER trial was supported by the Stichting Steunfonds Joodse GGZ and Sinai Centrum, The Netherlands (to KT). The authors like to thank Nick Lommerse for his assistance in retrieving the PROSPER clinical data, Adriaan Hoogendoorn for his input on the longitudinal statistical analyses and Niels de Joode, Tim van Balkom and Ton Schweigmann for their help in acquiring the healthy control imaging data.

## REFERENCES

1. Millan MJ, Agid Y, Brune M, Bullmore ET, Carter CS, Clayton NS, et al. (2012): Cognitive dysfunction in psychiatric disorders: characteristics, causes and the quest for improved therapy. Nat Rev Drug Discov 11: 141–68.

2. McTeague LM, Huemer J, Carreon DM, Jiang Y, Eickhoff SB, Etkin A (2017): Identification of Common Neural Circuit Disruptions in Cognitive Control Across Psychiatric Disorders. The American journal of psychiatry 174: 676–685.

3. Bullmore E, Sporns O (2009): Complex brain networks: graph theoretical analysis of structural and functional systems. Nature reviews Neuroscience 10: 186–98.

4. Lord LD, Stevner AB, Deco G, Kringelbach ML (2017): Understanding principles of integration and segregation using whole-brain computational connectomics: implications for neuropsychiatric disorders. Philosophical transactions Series A, Mathematical, physical, and engineering sciences 375. 10.1098/rsta.2016.0283

5. van den Heuvel MP, Sporns O (2019): A cross-disorder connectome landscape of brain dysconnectivity. Nature reviews Neuroscience 20: 435–446.

6. Spronk M, Keane BP, Ito T, Kulkarni K, Ji JL, Anticevic A, Cole MW (2021): A Whole-Brain and Cross-Diagnostic Perspective on Functional Brain Network Dysfunction. Cerebral Cortex 31: 547–561.

7. Sha Z, Wager TD, Mechelli A, He Y (2019): Common Dysfunction of Large-Scale Neurocognitive Networks Across Psychiatric Disorders. Biol Psychiatry 85: 379–388.

8. de Lange SC, Scholtens LH, Alzheimer’s Disease Neuroimaging I, van den Berg LH, Boks MP, Bozzali M, et al. (2019): Shared vulnerability for connectome alterations across psychiatric and neurological brain disorders. Nat Hum Behav 3: 988–998.

9. Hansen JY, Shafiei G, Vogel JW, Smart K, Bearden CE, Hoogman M, et al. (2022): Local molecular and global connectomic contributions to cross-disorder cortical abnormalities. Nat Commun 13: 4682.

10. Davis SW, Beynel L, Neacsiu AD, Luber BM, Bernhardt E, Lisanby SH, Strauman TJ (2023): Network-level dynamics underlying a combined rTMS and psychotherapy treatment for major depressive disorder: An exploratory network analysis. Int J Clin Health Psychol 23: 100382.

11. Szeszko PR, Yehuda R (2019): Magnetic resonance imaging predictors of psychotherapy treatment response in post-traumatic stress disorder: A role for the salience network. Psychiatry Res 277: 52–57.

12. Nord CL, Barrett LF, Lindquist KA, Ma Y, Marwood L, Satpute AB, Dalgleish T (2021): Neural effects of antidepressant medication and psychological treatments: a quantitative synthesis across three meta-analyses. Br J Psychiatry 219: 546–550.

13. Kim Y-K (2018): Treatment Resistance in Psychiatry: Risk Factors, Biology, and Management. Springer.

14. Gottlich M, Kramer UM, Kordon A, Hohagen F, Zurowski B (2015): Resting-state connectivity of the amygdala predicts response to cognitive behavioral therapy in obsessive compulsive disorder. Biological psychology 111: 100–9.

15. Zhutovsky P, Thomas RM, Olff M, van Rooij SJH, Kennis M, van Wingen GA, Geuze E (2019): Individual prediction of psychotherapy outcome in posttraumatic stress disorder using neuroimaging data. Translational psychiatry 9: 326.

16. Cyr M, Pagliaccio D, Yanes-Lukin P, Fontaine M, Rynn MA, Marsh R (2020): Altered network connectivity predicts response to cognitive-behavioral therapy in pediatric obsessive-compulsive disorder. Neuropsychopharmacology 45: 1232–1240.

17. Avissar M, Powell F, Ilieva I, Respino M, Gunning FM, Liston C, Dubin MJ (2017): Functional connectivity of the left DLPFC to striatum predicts treatment response of depression to TMS. Brain stimulation 10: 919–925.

18. Ge R, Downar J, Blumberger DM, Daskalakis ZJ, Vila-Rodriguez F (2020): Functional connectivity of the anterior cingulate cortex predicts treatment outcome for rTMS in treatment-resistant depression at 3-month follow-up. Brain stimulation 13: 206–214.

19. Colvonen PJ, Glassman LH, Crocker LD, Buttner MM, Orff H, Schiehser DM, et al. (2017): Pretreatment biomarkers predicting PTSD psychotherapy outcomes: A systematic review. Neuroscience and biobehavioral reviews 75: 140–156.

20. Cohen SE, Zantvoord JB, Wezenberg BN, Bockting CLH, van Wingen GA (2021): Magnetic resonance imaging for individual prediction of treatment response in major depressive disorder: a systematic review and meta-analysis. Transl Psychiatry 11: 1–10.

21. Gallen CL, D’Esposito M (2019): Brain Modularity: A Biomarker of Intervention-related Plasticity. Trends Cogn Sci 23: 293–304.

22. Insel TR, Cuthbert BN (2015): Medicine. Brain disorders? Precisely. Science 348: 499–500.

23. Snoek A, Beekman ATF, Dekker J, Aarts I, van Grootheest G, Blankers M, et al. (2020): A randomized controlled trial comparing the clinical efficacy and cost-effectiveness of eye movement desensitization and reprocessing (EMDR) and integrated EMDR-Dialectical Behavioural Therapy (DBT) in the treatment of patients with post-traumatic stress disorder and comorbid (Sub)clinical borderline personality disorder: study design. BMC Psychiatry 20: 396.

24. van den End A, Beekman ATF, Dekker J, Aarts I, Snoek A, Blankers M, et al. (2024): Trauma-focused and personality disorder treatment for posttraumatic stress disorder and comorbid cluster C personality disorder: a randomized clinical trial. Eur J Psychotraumatol 15: 2382652.

25. Wolf N, van Oppen P, Hoogendoorn AW, van den Heuvel OA, van Megen HJGM, Broekhuizen A, et al. (2024): Inference-Based Cognitive Behavioral Therapy versus Cognitive Behavioral Therapy for Obsessive-Compulsive Disorder: A Multisite Randomized Controlled Non-Inferiority Trial. Psychotherapy and Psychosomatics 1–15.

26. Fitzsimmons SMDD, Postma T, van Campen AD, Vriend C, Batelaan NM, van Oppen P, et al. (2024): TMS-induced plasticity improving cognitive control in OCD I: Clinical and neuroimaging outcomes from a randomised trial. Biol Psychiatry S0006-3223(24)01488–4.

27. van Balkom TD, Berendse HW, van der Werf YD, Twisk JWR, Peeters CFW, Hoogendoorn AW, et al. (2022): Effect of eight-week online cognitive training in Parkinson’s disease: A double-blind, randomized, controlled trial. Parkinsonism & Related Disorders 96: 80–87.

28. Pouwels PJW, Vriend C, Liu F, de Joode NT, Otaduy MCG, Pastorello B, et al. (2023): Global multi-center and multi-modal magnetic resonance imaging study of obsessive-compulsive disorder: Harmonization and monitoring of protocols in healthy volunteers and phantoms, 20220815th ed. Int J Methods Psychiatr Res 32: e1931.

29. Luppi AI, Gellersen HM, Liu Z-Q, Peattie ARD, Manktelow AE, Adapa R, et al. (2024): Systematic evaluation of fMRI data-processing pipelines for consistent functional connectomics. Nat Commun 15: 4745.

30. Ran Q, Jamoulle T, Schaeverbeke J, Meersmans K, Vandenberghe R, Dupont P (2020): Reproducibility of graph measures at the subject level using resting-state fMRI. Brain Behav 10: 2336–2351.

31. Tewarie P, van Dellen E, Hillebrand A, Stam CJ (2015): The minimum spanning tree: an unbiased method for brain network analysis. Neuroimage 104: 177–188.

32. Yeo BT, Krienen FM, Sepulcre J, Sabuncu MR, Lashkari D, Hollinshead M, et al. (2011): The organization of the human cerebral cortex estimated by intrinsic functional connectivity, 2011/06/10 ed. J Neurophysiol 106: 1125–65.

33. Mataix-Cols D, Cruz L, Nordsletten AE, Lenhard F, Isomura K, Simpson HB (2016): Towards an international expert consensus for defining treatment response, remission, recovery and relapse in obsessive-compulsive disorder. World Psychiatry 15: 80–81.

34. Chen G, Bürkner P-C, Taylor PA, Li Z, Yin L, Glen DR, et al. (2019): An integrative Bayesian approach to matrix-based analysis in neuroimaging. Human Brain Mapping 40: 4072–4090.

35. Chen G, Xiao Y, Taylor PA, Rajendra JK, Riggins T, Geng F, et al. (2019): Handling multiplicity in neuroimaging through Bayesian lenses with multilevel modeling. Neuroinformatics 17: 515–545.

36. Wilson DJ (2019): The harmonic mean p-value for combining dependent tests. Proceedings of the National Academy of Sciences 116: 1195–1200.

37. Arnemann KL, Chen AJ, Novakovic-Agopian T, Gratton C, Nomura EM, D’Esposito M (2015): Functional brain network modularity predicts response to cognitive training after brain injury. Neurology 84: 1568–74.

38. Doucet GE, Moser DA, Luber MJ, Leibu E, Frangou S (2020): Baseline brain structural and functional predictors of clinical outcome in the early course of schizophrenia. Mol Psychiatry 25: 863–872.

39. Gallen CL, Baniqued PL, Chapman SB, Aslan S, Keebler M, Didehbani N, D’Esposito M (2016): Modular Brain Network Organization Predicts Response to Cognitive Training in Older Adults. Plos One 11: e0169015.

40. Stern Y (2009): Cognitive reserve. Neuropsychologia 47: 2015–28.

