## supplementary material for "Treatment outcome is associated with pre-treatment connectome measures across psychiatric disorders - evidence for connectomic reserve?"

**SUPPLEMENTARY MATERIALS**

**Supplementary methods**

**Participants and interventions**

This project included participants from several completed clinical trials: ARRIBA (NCT03929081), TIPICCO (NCT03667807)(Fitzsimmons et al., 2024) and PROSPER (NCT03833531 / NCT03833453) (Snoek et al., 2020; van den End et al., 2024, 2021). Both ARRIBA and TIPICCO utilized the Yale-Brown Obsessive-Compulsive scale (YBOCS) before and after treatment as measure for overall OCD symptom severity. The PROSPER trials administered the clinician-administered PTSD interview for DSM-5 (CAPS-5) before and after the treatment. All participants from all trials received active treatment and provided written informed consent according to the declaration of Helsinki. All trials were approved by the Medical Ethical Committee of VU medical center. All participants of TIPICCO, but only a subset of the participants of the ARRIBA and PROSPER underwent MRI scanning. Only those participants with (at least) a baseline diffusion MRI (dMRI) and resting-state functional MRI (rsfMRI) and pre- and post-treatment clinical measures were included in the current analyses. Non treatment compliant individuals but with a post-treatment clinical assessment were still included (i.e. intention-to-treat sample). Below follows a description of the separate trials and treatments as well as the selected healthy control sample.

*ARRIBA*

Participants in ARRIBA were diagnosed with OCD and randomly assigned to Cognitive Behavioral therapy (CBT) consisting of either exposure and response prevention (ERP) therapy or Inference based cognitive behavioral therapy (I-CBT). Inclusion criteria were, being over the age of 18, have a primary diagnosis of obsessive-compulsive disorder (OCD) according to the DSM-5 with moderate-to-severe OCD symptoms as evidence by a Yale-Brown Obsessive-Compulsive Scale (Y-BOCS) score of at least 16, not being on any psychotropic medication or on a stable dose for at least 12 weeks prior to the study and during the study and not having received any cognitive behavioral therapy in the 6 months before the study. Participants were excluded in case of a psychotic disorder, organic mental disorder, substance abuse disorder, mental retardation or insufficient comprehension of the Dutch language. Trial participants were randomized to either cognitive behavioral therapy with exposure and response prevention therapy (CBT-ERP) or inference-based cognitive behavioral therapy (I-CBT). Both treatments were protocolized into 20 weekly sessions of 45 minutes and delivered by trained psychologists. A subset of the participants underwent MRI at Amsterdam UMC before and after treatment. Full treatment protocol and main clinical results on the full sample of N=197 are described elsewhere (Wolf et al., 2024). This Dutch multicenter trial was prospectively registered at ClinicalTrials.gov (NCT03929081).

*TIPICCO*

TIPICCO participants were also diagnosed with OCD and received ERP in combination with repetitive transcranial magnetic stimulation (rTMS) over the dorsolateral prefrontal cortex (DLPFC), presupplementary motor area (pre-SMA) or vertex. Participants had to be aged between 18-65, have a primary diagnosis of OCD according to the DSM-5, score at least 16 on the YBOCS, underwent previous treatment with 8 or more sessions of CBT or ERP and 12 or more weeks of treatment with Serotonin reuptake inhibitors or being medication-naive. Participants had to keep a stable dose of medication throughout the intervention period. Exclusion criteria were Tourette’s disorder, schizophrenia, bipolar disorder, active suicidal ideation, prior exposure to transcranial magnetic stimulation (rTMS) or contraindications for MRI or rTMS. Participants were randomized to eight weeks, twice weekly (16 sessions) CBT-ERP in combination with either (1) rTMS to the left dorsolateral prefrontal cortex, (2) rTMS to the pre-supplementary motor area (preSMA) or (3) rTMS to the vertex (as a control condition). participants in the DLPFC or pre-SMA rTMS conditions received 10 Hz rTMS at 110% resting motor threshold, while participants in the vertex condition received 10-Hz at 60% resting motor threshold. All participants received 3000 pulses per session (30 x 10 second trains with 30-second intertrain intervals). See Fitzsimmons for details on selection and personalization of the rTMS targets. All CBT-ERP sessions started within 10 minutes after rTMS and lasted for 60 minutes and were provided by trained psychotherapists. This Dutch trial was prospectively registered at ClinicalTrials.gov (NCT03667807). Full treatment protocol and main clinical results on the full sample of N=66 are described elsewhere (Fitzsimmons. 2024).

*PROSPER-B*

Participants were diagnosed with PTSD and (1) a comorbid (subclinical) borderline personality disorder. PROSPER-B participants were randomized to eye movement desensitization and reprocessing (EMDR) therapy or integrated EMDR and dialectical behavioral therapy (DBT) (Snoek et al., 2020). Eligibility criteria consisted of participants being 18-65 years of age, had a primary diagnosis of post-traumatic stress disorder (PTSD) and at least four symptoms of a borderline personality disorder (BPD) according to DSM-5 criteria. In case of psychotropic medication, participants had to be on a stable dose for at least three weeks. Exclusion criteria were other concurrent psychological treatment, treatment-interfering comorbidities (current psychosis, paranoid, schizotypal, histrionic, narcissistic, or antisocial personality disorder, substance disorders, BMI< 17 or IQ<70) or insufficient comprehension of the Dutch language. participants were randomized to either stand-alone eye movement desensitization and reprocessing (EMDR) therapy or EMDR in combination with dialectical behavioral therapy (DBT). Participants underwent 12-18 weekly EMDR sessions of each 60 minutes. DBT was provided concurrently for 54 weeks to the EMDR+DBT condition. Treatment starts with six weekly 45 minute individual DBT sessions with thereafter individual psychotherapy and group skills training that consists of 48 weekly group sessions of 150 minutes. Full treatment protocol has been described elsewhere (Snoek et al. 2020). A manuscript on the main clinical results on the full sample of N=124 is currently under revision. The trial was prospectively registered at ClinicalTrials.gov (NCT03833453).

*PROSPER-C*

Participants were diagnosed with PTSD and a comorbid cluster C personality (PROSPER-C) and received either imagery rescripting alone or imagery rescripting + schema therapy (van den End et al., 2021). Eligibility criteria consisted of participants being 18-65 years of age, had a primary diagnosis of post-traumatic stress disorder (PTSD) according to DSM-5 criteria and exhibit symptoms of a cluster C personality disorder (PD): at least three symptoms of avoidant PD, four symptoms of dependent PD or three symptoms of obsessive-compulsive PD. In case of psychotropic medication, participants had to be on a stable dose for at least three weeks. Exclusion criteria were a current psychosis, severe aggression, treatment-interfering substance or eating disorders, somatic problems, mental retardation or insufficient comprehension of the Dutch language. Participants were randomized to PTSD-focused imagery rescripting (ImRs) or ImRs in combination with group-based schema therapy (ST). Participants in the ImRs condition received 12-18 weekly sessions of 75 minutes in a span of maximally six months. ST consisted of four 45-min individual pre-treatment sessions and 40 weekly group sessions of 90 minutes. Sessions were administered by therapists with at least three years of experience. Full treatment protocol and main clinical results on the full sample of N=130 are described elsewhere(van den End et al., 2024, 2021). The trial was prospectively registered at ClinicalTrials.gov (NCT03833531).

*Healthy controls*

A healthy control sample was selected from a pool of healthy controls that was scanned on the same MRI scanner as part of the ARRIBA, TIPICCO and PROSPER-B/C trials, supplemented with healthy controls from the COGTIPS trial (van Balkom et al., 2022) and global OCD study (Pouwels et al., 2023) All healthy controls were free from any current psychopathology, psychotropic medication or neurological disorders. Healthy controls in the age range 18-65 were matched to the patient sample on age and sex (see Methods - data analyses).

**Image acquisition and preprocessing**

All participants were scanned on the same GE Discovery MR750 3T (General Electric, Milwaukee, U.S) using a 32-channel head coil with harmonized sequences. We acquired T1-weighted structural magnetization-prepared rapid acquisition gradient-echo (MPRAGE; TR = 6.9 ms; TI = 900 ms; TE = 3.0 ms; 256x256 matrix; 1 mm3 isotropic resolution; 168 sections). The ten minutes eyes-closed resting-state fMRI scans consisted of a gradient echo-planar imaging (TR = 2.2 s; TE = 26 ms; 64x64 matrix; field of view 21.1cm; flip angle = 80°) with 42 ascending slices per volume (3.3 x 3.3 mm in-plane resolution; slice thickness = 3.0 mm; interslice gap = 0.3 mm). DMRI multi-shell images consisted of 73 diffusion-weighted directions (25 b1000, 24 b2000, and 24 b3000 s/mm^2^) and seven interleaved non-diffusion-weighted volumes (b0 s/mm^2^). Fieldmaps with opposite phase-encoding directions were acquired for both the rsfMRI and dMRI. RsfMRI data were pre-processed using fMRIprep v21.0.1 ([https://fmriprep.org](https://fmriprep.org/en/stable/)) (see boilerplate at the end of the supplements), consisting among others of realignment (and motion parameters calculation), slice-timing, susceptibility induced distortion correction using the field maps and co-registration to the T1 image. We removed the first three non-steady-state volumes and smoothed the functional scan with a 6 mm FWHM kernel using FSL SUSAN, prior to simultaneous nuisance regression and temporal filtering (0.009 – 0.08 Hz) using a ‘24HMP8PhysSpikeRegGS’ pipeline (Satterthwaite et al., 2013): 24 head motion parameters (HMP; translations, rotations, their temporal derivatives and quadratic terms), 8 physiological regressors from the white matter and CSF (8Phys; mean, temporal derivates and quadratic terms, spike regressors (SpikeReg) based on a frame-to-frame displacement (FD) > 0.5 mm and the global signal (GS). Scans with a mean FD>0.5 mm or incomplete coverage of the atlas regions were excluded. DMRI scans were denoised using dwidenoise in MRTRIX3 and subsequently corrected for eddy current, susceptibility-induced distortions, and within- and between-volume motion using FSL eddy. The resulting images were visually inspected for residual (motion) artifacts and if necessary excluded. We performed multi-shell anatomically-constrained (probabilistic) tractography in MRtrix3 on the preprocessed dMRI with 50 million seeds from the gray/white matter boundary after generating a five-tissue-type (5TT) segmentation based on Hybrid Surface and Volume segmentation (HSVS) using FreeSurfer (v7.3.2) output. We additionally applied SIFT2 to improve the accuracy of the reconstructed fibers and reduce false positives in the tractogram (McColgan et al., 2018; Smith et al., 2015). Scans with incomplete coverage of the atlas regions were excluded.

**Connectome construction**

We used 300 cortical areas derived from the Schaefer 300P7N atlas and 14 individually segmented subcortical areas with FreeSurfer 7.3.2 to extract time series from the preprocessed and denoised functional image. Based on previous evaluations of different fMRI processing pipelines for consistent and reproducible network reconstruction (Luppi et al., 2024; Ran et al., 2020), we used Pearson correlations to cross-correlate the time series, absolutized negative correlations and applied Orthogonal Minimal Spanning Trees (OMST) thresholding - a data-driven filtering method that balances network efficiency and wiring cost (Dimitriadis et al., 2017) to construct a 314x314 weighted functional connectome. The dMRI-derived tractogram was converted to a weighted structural connectome (with streamline count as edge weight) using the same 300P7N Schaefer cortical atlas and 14 subcortical areas and thresholded using OMST. To construct the multilayer connectome, we first applied a minimum spanning tree (MST) algorithm to the structural and functional connectomes to determine their network backbone. This backbone is a binarized version made up of the strongest links of the weighted version that connect all the nodes in the network without forming loops. Connections between the same nodes in the structural and functional layers of the multiplex (i.e. interlayer links) are also represented with a weight of 1.

**Network measures**

For the structural and functional connectomes we calculated: global efficiency and average participation coefficient PC, both representing integration, modularity (segregation) and small-worldness (balance of integration and segregation). Modularity (quality index) and (average) PC were primarily calculated using the Yeo 7 network parcellation, (Yeo et al., 2011) but also using a participant-specific data-driven community detection approach (generalized Louvain heuristic). The structural and functional connectomes were additionally reduced to an 8x8 ‘system’ network consisting of the seven Yeo cortical networks (Default mode network [DMN], Frontoparietal Network [FPN], Dorsal attention Network [DAN], Ventral Attention Network [VAN], Somatomotor Network [SMN], limbic and Visual Network) and subcortical brain areas by averaging the edge weights of each of the nodes that connect these ‘systems’ and used as input for a matrix-based Bayesian analysis to assess the between system connectivity strength (see statistical analyses in the main text). We also calculated the average PC of the nodes of each of the eight systems as a measure of the diversity of the connections within versus across systems. Because ‘multilayer network analysis’ is still an evolving field, not all single layer network measures have a suitable multilayer counterpart. From the multilayer network, we calculated the average eccentricity and eigenvector centrality for the whole network and the eight systems. Nodal eccentricity represents the maximum distance between the node and all others. The average eccentricity therefore provides a measure for (dis)integration as it represents how far information has to travel to cross the network. Eigenvector centrality is a measure for how influential a node is for network communication. Higher average eigenvector centrality represents a more integrated network structure with many central nodes. See for the mathematical equations below.

| Description of the network measures | | | | |
| --- | --- | --- | --- | --- |
| Measure | Modality | level | Description | Formula |
| Global efficiency (Eglob) | Functional  structural | global | efficiency of information exchange in a network. Defined as the inverse of the characteristic path length | $Eglob= \frac{1}{N(N-1)}\sum_{i\neq j} d_{ij}$  $d_{ij}=shortest path nodes i to j$ |
| Modularity (Q) | Functional  Structural | global | Degree to which a network can be divided into distinct communities or modules. | $Q= \frac{1}{2m}\sum_{ij} \lfloor A_{ij}- \frac{kikj}{2m}\rfloor d(ci,cj)$  $A_{ij}=adjaceny matrix$  $k_{i}$ / $k_{j}$ = degree of node I and j  d = 1 if nodes in same module, 0 otherwise |
| Average Participation coefficient (PC) | Functional  Structural | global  system | Assesses how evenly a node's links are distributed across different modules in the network. Higher PC means more equal distribution of connections across modules | $Pi=1- \sum_{s=1}^{M} \left( \frac{k_{i}^{s}}{k_{i}} \right)^{2}$  $k_{i}^{s}=$ number of connections of node i in module s  $k_{i}=$ total degree of node i. |
| Small-Worldness (Sigma) | Functional  Structural | global | A global measure of a network's ratio of clustering to path length relative to a random network. | $Sigma= \frac{C/Crand}{L/Lrand}$  $C=clustering coefficient$  $L=characteristic path length$  $rand=random network$ |
| Average eccentricity | multilayer | global  system | average of the maximum distances (shortest paths) between each node and all other nodes. | $e\left( i \right)=max(d_{ij})$  $d_{ij}=shortest path nodes i to j$ |
| Average Eigenvector centrality | multilayer | global  system | Measures a node's influence based on its own connections and the connections of its neighbors. | $EC\left( i \right)= \frac{1}{}\sum_{j} A_{ij}EC(j)$  $A_{ij}=adjaceny matrix$  $=largest eigenvalue$ |

**Statistical analysis**

*Advantages of Bayesian multilevel approaches for brain measures.*

The Bayesian approach simultaneously associated treatment outcome with all between system connections in one statistical model thereby (1) taking into account the inherent shared information across all matrix entries (i.e. all derived from the same brain), (2) mitigating the need for multiple comparison correction across models and (3) being more respectful of the continuous nature of biological measures. It additionally better controls magnitude and sign errors, and stimulates full reporting of the results thereby improving transparency and reproducibility.(Chen et al., 2019; Taylor et al., 2023)

*Deviations from pre-registration plan for pre-to-post treatment change analyses*

In the pre-registration plan we stated that we would use univariate mixed model analyses with the network measures at post-treatment as dependent variable and treatment outcome expressed as percentage change or responder/non-responder as independent variable and the pre-treatment value of the network measures, along with age and sex as covariates to investigate how single-layer and multilayer topology of the network changes with successful treatment. Trial or treatment would be added as random intercept in separate sensitivity analyses. During the analyses we realized that this model is unsuited to adequately capture the associations between changes in clinical symptoms or response status and changes in network measures but instead determines whether there is a significant association between *post-treatment* network measures and the change in clinical symptoms/response status while adjusting for the pre-treatment value of the network measures (and age, sex and trial/treatment). For the percentage change models we therefore z-transformed the individual-centered pre-treatment and post-treatment clinical severity scores (to adjust for differences in scales between trials) and subsequently associated these values with the pre-to-post treatment global network measures in a hierarchical linear regression model with ‘participant’ as a fixed effect. Because participant is used as fixed effect, additionally adjusting for age and sex does not add to the model. These models are equivalent to a repeated measures correlation (rmcorr (Bakdash and Marusich, 2017)) when the same scale is used. For the responder vs non-responder analyses we used mixed model analyses with a random intercept for participant, the network measure as dependent variable and an interaction term for time and response status (along with their main effects), while adjusting for the pre-treatment value of the network measure, age and sex. Trial or treatment were added as additional covariates in separate sensitivity analyses. For both sets of analyses we used a statistical threshold of p < 0.05. Code examples are provided on the author’s github page: [github.com/chrisvriend/core-multi](http://github.com/chrisvriend/core-multi)

*Additional preregistered Pre-to-post treatment*

Because the Bayesian multilevel modeling framework does not support within-subject designs, we first calculated the change in system connectivity from pre-to-post treatment, change in system PC (single layers), eccentricity and eigenvector centrality (multilayer) and associated these values with percentage change and compared them between responders and non-responders. NBS was performed on the post-treatment – pre-treatment difference matrix and associated with improvement in symptoms.

**Supplementary Results**

Flowchart

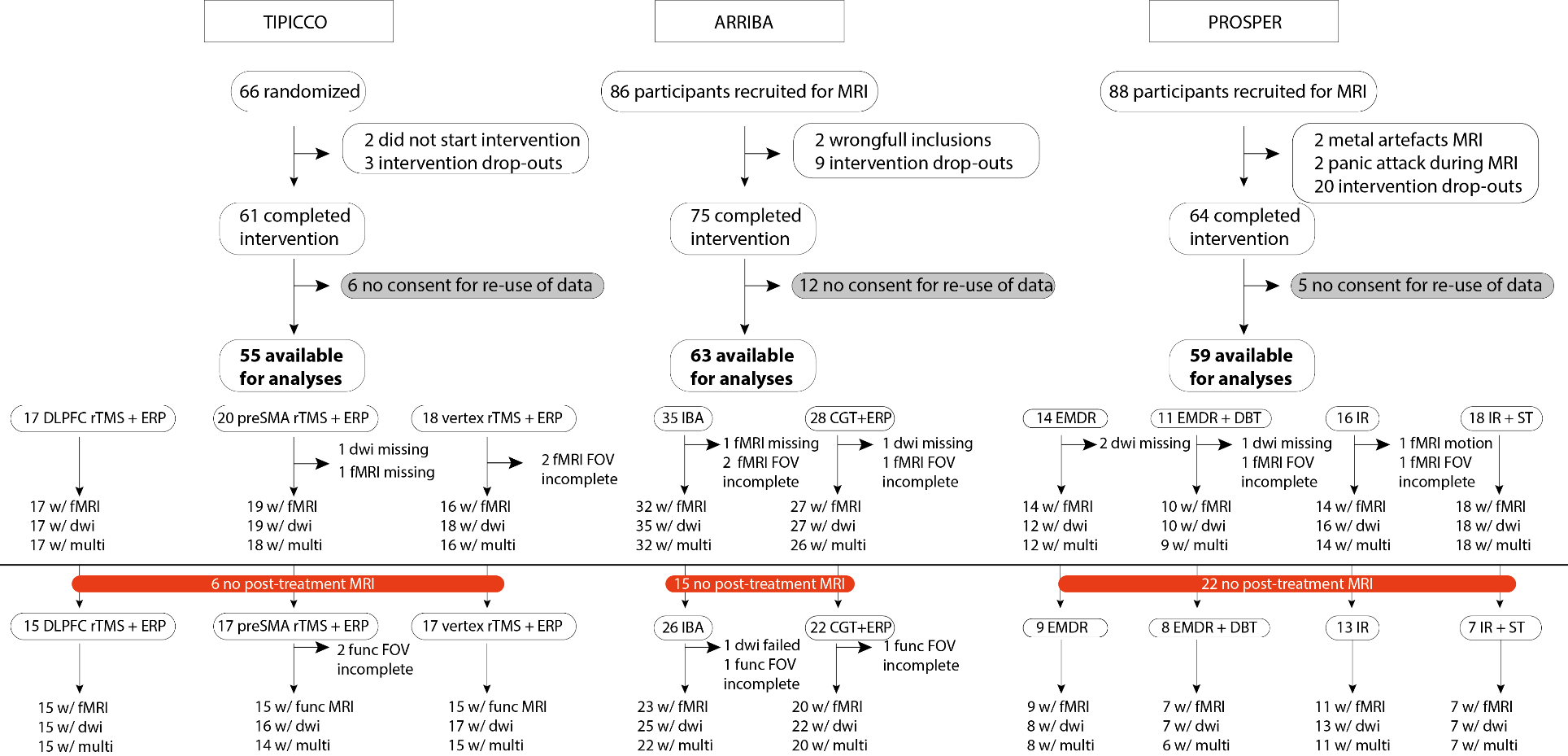

**Supplementary figure 1 – Flowchart**

Abbreviations: CGT-ERP = cognitive behavioral therapy (CBT) with exposure and response prevention (ERP) therapy, IBA = inference based approach, DLPFC rTMS + ERP = repetitive Transcranial Magnetic Stimulation (rTMS) to the dorsolateral prefrontal cortex in combination with ERP, preSMA rTMS + ERP = rTMS to the pre-supplementary motor area in combination with ERP, vertex rTMS + ERP = rTMS to the vertex in combination with ERP, EMDR = eye-movement desensitization and reprocessing, EMDR+DBT = EMDR and dialectical behavioral therapy, IR = imagery rescripting therapy, IR+ST = imagery rescripting and schema focused therapy.

| **Supplementary Table 1 – Demographic and clinical characteristics of pre-post treatment sample** | | | | | | | | | | |
| --- | --- | --- | --- | --- | --- | --- | --- | --- | --- | --- |
| **Sample** | **Treatment** | **N** | **Age (yrs)** | **T0 sev.#** | **T1 sev.#** | **% improvement** | **Sex**  **(F/M)** | **Responder**  **(no/yes)** | **Med (no/yes)** | **FD**  **fMRI** |
| **Total** |  | 134 | 36.7 ± 11.7 | 31 ± 10.6 | 17.5 ± 11 | 43.3 ± 27 | 80/54 | 49/85 | 66/67* | 0.13 ± 0.07 |
| ARRIBA | CBT-ERP | 22 | 30.9 ± 10 | 24.3 ± 3.6 | 10.2 ± 6 | 57.5 ± 26.7 | 11/11 | 6/16 | 13/9 | 0.12 ± 0.05 |
|  | I-CBT | 26 | 34.6 ± 10.3 | 24.3 ± 4.5 | 16.5 ± 6.9 | 32.9 ± 22.1 | 13/13 | 12/14 | 14/12 | 0.13 ± 0.05 |
| TIPICCO | DLPFC | 15 | 39.9 ± 11.9 | 28.9 ± 6 | 18.1 ± 7.5 | 37.4 ± 20.4 | 9/6 | 6/9 | 5/10 | 0.15 ± 0.09 |
|  | preSMA | 17 | 30.9 ± 13.2 | 29 ± 3.6 | 18.7 ± 8.2 | 36.2 ± 26 | 9/8 | 9/8 | 5/12 | 0.14 ± 0.07 |
|  | vertex | 17 | 40.3 ± 11.9 | 26.6 ± 4.1 | 15.5 ± 4.9 | 40.7 ± 20.1 | 12/5 | 6/11 | 9/8 | 0.12 ± 0.05 |
| PROSPER-B | EMDR | 8 | 40.2 ± 6.5 | 43.5 ± 12 | 24.6 ± 13.8 | 43.6 ± 26.7 | 6/2 | 3/5 | 5/3 | 0.12 ± 0.04 |
|  | EMDR+DBT | 9 | 37.8 ± 12.8 | 45.4 ± 9.4 | 30 ± 15.8 | 35.6 ± 32.2 | 8/1 | 3/6 | 4/5 | 0.12 ± 0.05 |
| PROSPER-C | IR | 13 | 43 ± 11.1 | 37.5 ± 12.1 | 15.6 ± 15.7 | 59.3 ± 32.9 | 9/4 | 2/11 | 6/6 | 0.16 ± 0.13 |
|  | IR+SFT | 7 | 43.7 ± 10.2 | 51.9 ± 7.4 | 24.6 ± 18.9 | 54.3 ± 33.7 | 3/4 | 2/5 | 5/2 | 0.13 ± 0.04 |
| Scores are presented as mean ± standard deviation, unless otherwise indicated. # score represents the severity on the Yale-Brown Obsessive-compulsive Scale (Y-BOCS) for the ARRIBA and TIPICCO trials, and the clinician-administered PTSD scale for DSM-5 (CAPS-5) for the PROSPER trials. Medication is operationalized as using selective serotonin reuptake inhibitors (SSRI’s), serotonin-norepinephrine reuptake inhibitors (SNRI), tricyclic antidepressants (TCAs), antipsychotics or mood stabilizers at the time of the T0 MRI scan. * data was missing for one case. Abbreviations: CBT-ERP = cognitive behavioral therapy (CBT) with exposure and response prevention (ERP) therapy, I-CBT = inference based CBT, DLPFC = repetitive Transcranial Magnetic Stimulation (rTMS) to the dorsolateral prefrontal cortex in combination with ERP, preSMA = rTMS to the pre-supplementary motor area in combination with ERP, vertex = rTMS to the vertex in combination with ERP, EMDR = eye-movement desensitization and reprocessing, EMDR+DBT = EMDR and dialectical behavioral therapy, IR = imagery rescripting therapy, IR+SFT = imagery rescripting and schema focused therapy. | | | | | | | | | | |

Case-control analyses

Case-control analyses showed on the global level significantly lower global efficiency (B[SE]=-3.53[1.2], P=0.004), modularity (B[SE]=-4.6[0.6], P<0.001) and small-worldness (B[SE]=-14.7[5.7], P=0.01) and higher average PC (B[SE]=3.5[0.4], P<0.001) in the patient population for the functional but not structural connectome and higher multilayer eccentricity (B[SE]=0.7[0.3], P=0.02; see Figure 1 and supplementary Table 2-3). This suggests that cases have suboptimal integrative and segregative properties of the functional connectome and lower multilayer integrative capacity. These results were also evident at the system level (see supplementary Figures 2-3).

| **Supplementary Table 2 – case-control analyses of global connectome measures** | | | | | | | | |
| --- | --- | --- | --- | --- | --- | --- | --- | --- |
|  | **crude model** | | | **adjusted model*** | | | **LOSO** | |
|  | B [SE] | 95% CI | P | B [SE] | 95% CI | P | Trial  (harm.P) | Treatment  (harm.P) |
| **FUNCTIONAL CONNECTOME** | | | | | | | | |
| Eglob (x 10^-3^) | -3.363 [1.230] | -5.78 \| -0.94 | 0.007 | -3.294 [1.240] | -5.73 \| -0.86 | **0.008** | <0.001 | <0.001 |
| Q_yeo_ (x 10^-2^) | -4.500 [0.540] | -5.56 \| -3.44 | 0.000 | -4.561 [0.530] | -5.6 \| -3.52 | **<0.001** | <0.001 | <0.001 |
| PC_yeo_ (x 10^-2^) | 3.416 [0.430] | 2.58 \| 4.25 | 0.000 | 3.503 [0.410] | 2.69 \| 4.31 | **<0.001** | <0.001 | <0.001 |
| Small-worldness (x 10^-2^) | -15.088 [5.750] | -26.4 \| -3.78 | 0.009 | -15.276 [5.730] | -26.56 \| -3.99 | **0.008** | <0.001 | <0.001 |
| **STRUCTURAL CONNECTOME** | | | | | | | | |
| Eglob (x 10^-4^) | -7.698 [6.330] | -20.15 \| 4.76 | 0.225 | -7.374 [6.390] | -19.94 \| 5.19 | 0.249 | 0.24 | 0.145 |
| Q_yeo_ (x 10^-2^) | -0.001 [0.260] | -0.52 \| 0.52 | 0.997 | 0.045 [0.260] | -0.47 \| 0.56 | 0.863 | 0.743 | 1.00 |
| PC_yeo_ (x 10^-2^) | -0.003 [0.120] | -0.23 \| 0.23 | 0.978 | -0.028 [0.120] | -0.26 \| 0.2 | 0.808 | 0.968 | 1.00 |
| Small-worldness (x 10^-2^) | 7.721 [18.820] | -29.32 \| 44.76 | 0.682 | 14.409 [18.450] | -21.9 \| 50.72 | 0.436 | 0.631 | 0.688 |
| **MULTILAYER CONNECTOME** | | | | | | | | |
| EC (x 10^-2^) | -0.321 [0.300] | -0.9 \| 0.26 | 0.278 | -0.264 [0.300] | -0.85 \| 0.32 | 0.373 | 0.497 | 0.527 |
| Ecc (x 10^-2^) | 0.613 [0.290] | 0.04 \| 1.18 | 0.035 | 0.707 [0.290] | 0.14 \| 1.27 | **0.014** | <0.001 | <0.001 |
| *Corrected for age and sex. Abbreviations: LOSO = leave-one-sample-out – either one of the trials or one of the treatments. Harm. P = harmonized P-value, CI = confidence interval, Eglob = global efficiency, Q = modularity quality index, PC = participation coefficient, EC = eigenvector centrality, Ecc = eccentricity. | | | | | | | | |

| **Supplementary Table 3 – case-control analyses of global connectome measures - adjusting for treatment or trial** | | | | | | |
| --- | --- | --- | --- | --- | --- | --- |
|  | **Adjusted for treatment** | | | **Adjusted for trial** | | |
|  | B [SE] | 95% CI | P | B [SE] | 95% CI | P |
| **FUNCTIONAL CONNECTOME** | | | | | | |
| Eglob (x 10^-3^) | -3.300 [1.200] | -5.9 \| -0.8 | 0.008 | -3.300 [1.200] | -5.9 \| -0.7 | 0.008 |
| Q_yeo_ (x 10^-2^) | -4.500 [0.900] | -5.9 \| -3 | 0.066 | -4.300 [1.300] | -6.5 \| -2.1 | 0.089 |
| PC_yeo_ (x 10^-2^) | 3.500 [0.600] | 2.4 \| 4.5 | 0.079 | 3.400 [0.800] | 2.1 \| 4.6 | 0.088 |
| Small-worldness (x 10^-2^) | -14.800 [9.700] | -31.7 \| 2.2 | 0.258 | -15.300 [5.700] | -27.3 \| -3.1 | 0.008 |
| **STRUCTURAL CONNECTOME** | | | | | | |
| Eglob (x 10^-4^) | -7.400 [6.400] | -19.8 \| 5.1 | 0.249 | -7.400 [6.400] | -20 \| 5.2 | 0.249 |
| Q_yeo_ (x 10^-2^) | 0.100 [0.300] | -0.6 \| 0.7 | 0.908 | 0.000 [0.600] | -1 \| 1 | 0.987 |
| PC_yeo_ (x 10^-2^) | -0.000 [0.100] | -0.3 \| 0.2 | 0.808 | -0.000 [0.100] | -0.3 \| 0.2 | 0.808 |
| Small-worldness (x 10^-2^) | 14.400 [18.500] | -21.6 \| 50.5 | 0.436 | 14.200 [19.600] | -27.5 \| 53.6 | 0.650 |
| **MULTILAYER CONNECTOME** | | | | | | |
| EC (x 10^-2^) | -0.300 [0.300] | -0.8 \| 0.3 | 0.669 | -0.200 [0.500] | -1.1 \| 0.6 | 0.746 |
| Ecc (x 10^-2^) | 0.700 [0.600] | 0.1 \| 1.3 | 0.397 | 0.700 [0.400] | 0 \| 1.4 | 0.371 |

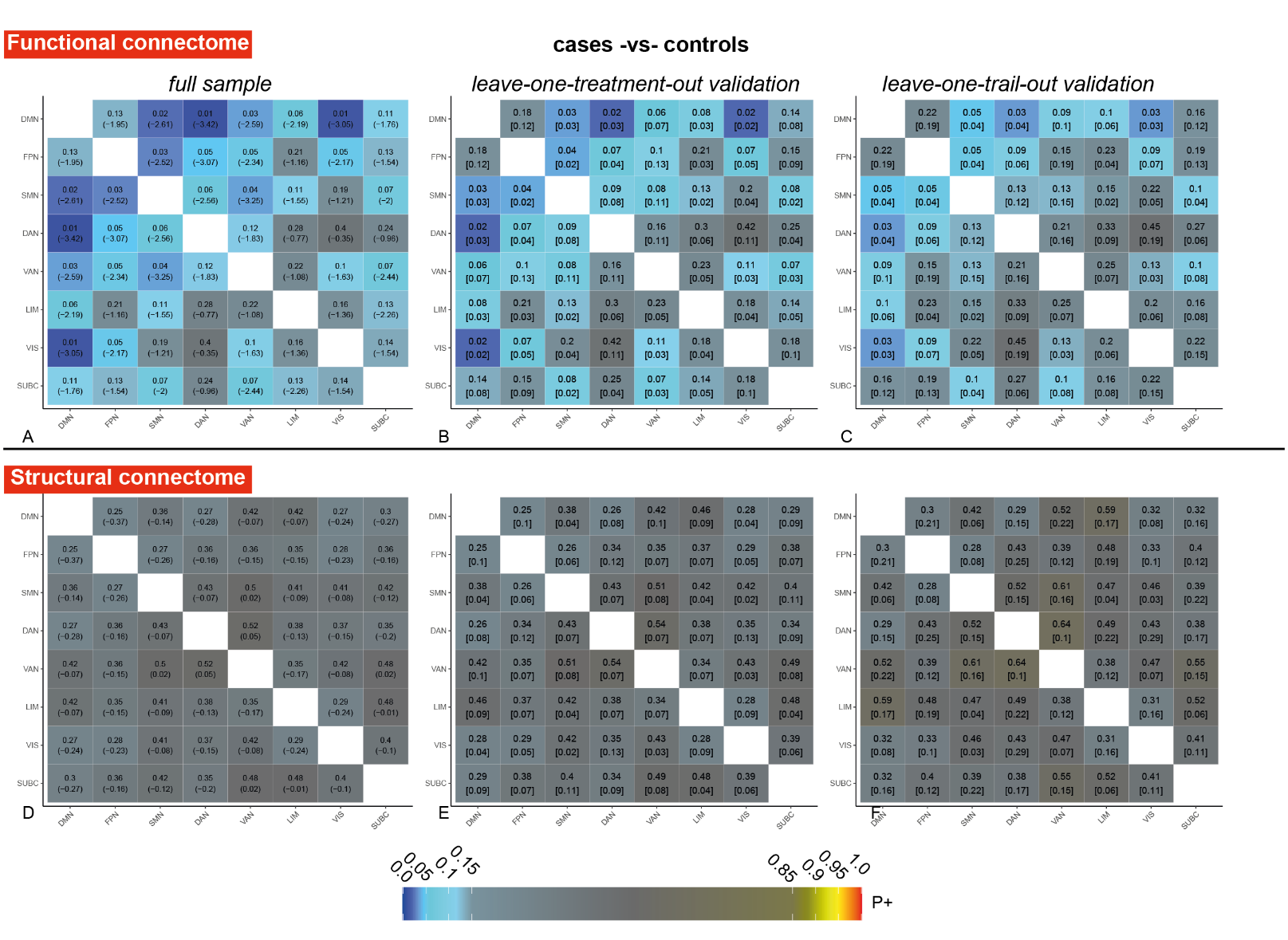

**Supplementary Figure 2 – Case-control system connectivity analyses.** Matrix-based Bayesian analyses showing case-control connectivity differences between 8 systems of the functional (a-c) and structural (d-f) connectome. Each square in the matrix represents the connectivity between two systems (matrix is symmetric). The color (dark-blue to light blue / yellow to red) represents the level of credibility of evidence according to the positive posterior probability (P+). The top values in each of the squares for the full sample (a/d) represent the P+ value and the bottom value between brackets the distance of the mode of the posterior probability plot to the zero-effect line (as measure for effect size and direction of effect). Top values in the leave-one-treatment-out (b/e) and leave-one-trial-out (c/f) validation matrices represent the median P+ values across folds. The [bottom value] represents the interquartile range across folds. Results indicate widespread and generally robust lower functional but not structural connectivity between systems in psychiatric cases compared with healthy controls. Abbreviations: DMN = default mode network, FPN = frontoparietal network, SMN = somatomotor network, DAN = dorsal attention network, VAN = ventral attention network, LIM = limbic network, VIS = visual network. SUBC = subcortical structures

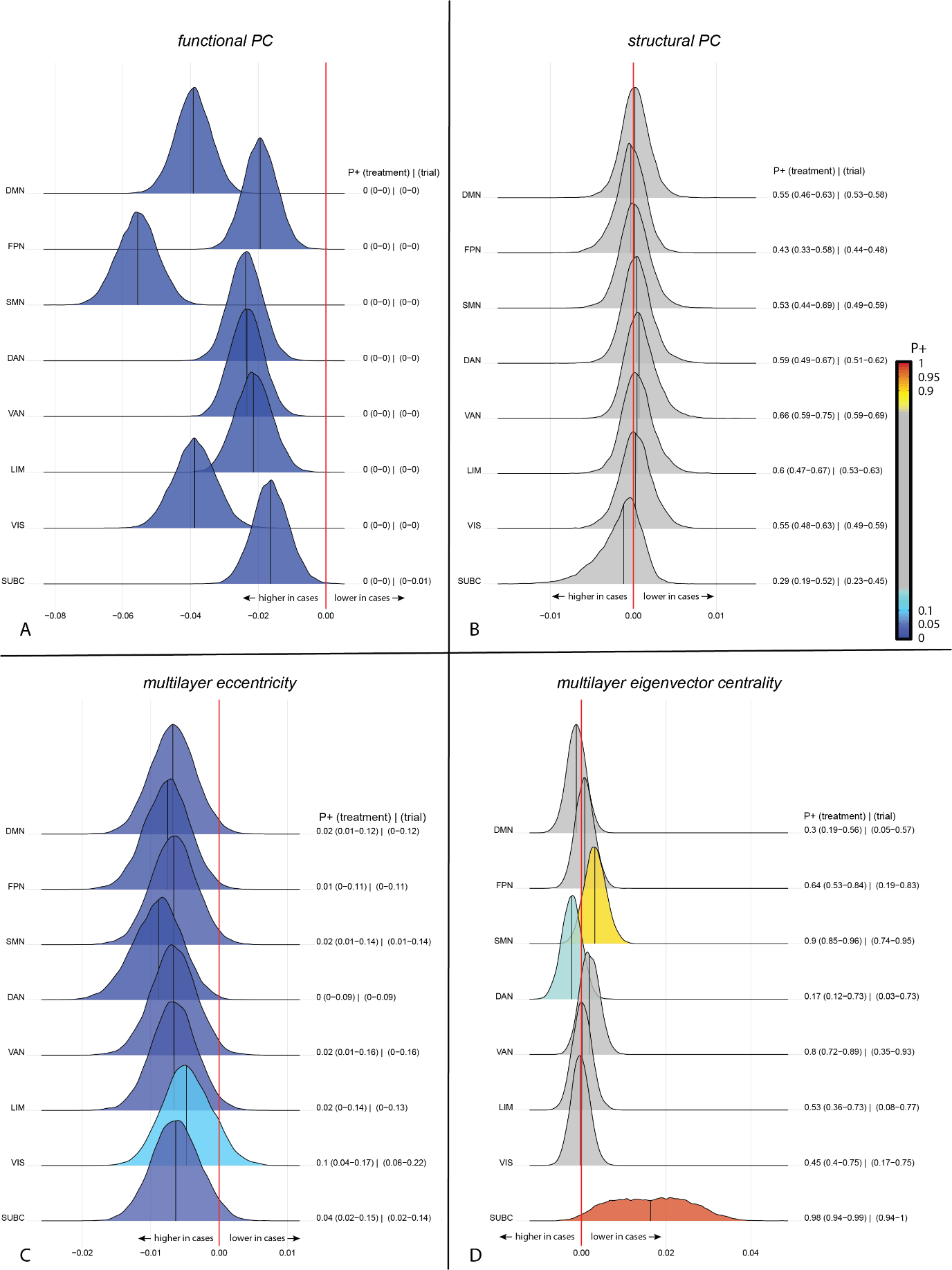

**Supplementary Figure 3 – case-control differences in single layer participation coefficient (PC), multilayer eccentricity and eigenvector centrality.** The posterior distribution communicates the credibility of an effect. Positive posterior probabilities (*P+*) are shown next to each distribution and color coded. The values between brackets indicate the range of P+ values across leave-one-treatment-out, or leave-one-trial-out folds to show the robustness of the results. *P+* values ≥0.90 indicate moderate to very high credibility for a positive effect, P+ ≤0.10 indicate moderate to very high credibility for a negative effect. The meaning of the direction of effects are shown next to the red zero-effect line. There was credible evidence for (a) higher functional participation coefficient (PC), but no differences in (b) structural PC in psychiatric cases compared with healthy controls across all eight systems. (c) multilayer eccentricity was also higher in cases, while (d) multilayer eigenvector centrality was lower in the SMN and subcortical areas. abbreviations: PC = participation coefficient, DMN = default mode network, FPN = frontoparietal network, SMN = somatomotor network, DAN = dorsal attention network, VAN = ventral attention network, LIM = limbic network, VIS = visual network. SUBC = subcortical structures.

Treatment outcome and pre-treatment network measures

| **Supplementary Table 4 – pre-treatment global connectome measures and treatment response** | | | | | | | | |
| --- | --- | --- | --- | --- | --- | --- | --- | --- |
|  | **crude model** | | | **adjusted model*** | | | **LOSO** | |
| **% CHANGE** | B [SE] | 95% CI | P | B [SE] | 95% CI | P | Trial  (harm.P) | Treatment  (harm. P) |
| **FUNCTIONAL CONNECTOME** | | | | | | | | |
| Eglob (x 10^-3^) | -4.700 [2.600] | -9.8 \| 0.4 | 0.068 | -4.500 [2.600] | -9.6 \| 0.5 | 0.077 | 0.033 | 0.001 |
| Q_yeo_ (x 10^-2^) | 2.400 [1.400] | -0.4 \| 5.2 | 0.097 | 2.500 [1.400] | -0.2 \| 5.2 | 0.074 | 0.021 | 0.001 |
| PC_yeo_ (x 10^-2^) | -1.900 [1.100] | -4.1 \| 0.3 | 0.084 | -2.000 [1.100] | -4.1 \| 0.1 | 0.059 | 0.013 | <0.001 |
| Small-worldness (x 10^-2^) | 24.400 [12.600] | -0.5 \| 49.4 | 0.055 | 25.400 [12.400] | 0.9 \| 49.9 | **0.042** | 0.009 | <0.001 |
| **STRUCTURAL CONNECTOME** | | | | | | | | |
| Eglob (x 10^-4^) | 11.000 [13.900] | -16.3 \| 38.4 | 0.428 | 11.000 [13.900] | -16.5 \| 38.5 | 0.433 | 0.49 | 0.655 |
| Q_yeo_ (x 10^-2^) | 0.300 [0.500] | -0.8 \| 1.4 | 0.601 | 0.200 [0.500] | -0.8 \| 1.3 | 0.652 | 0.827 | 0.898 |
| PC_yeo_ (x 10^-2^) | 0.100 [0.200] | -0.3 \| 0.5 | 0.632 | 0.100 [0.200] | -0.3 \| 0.6 | 0.565 | 0.828 | 0.922 |
| Small-worldness (x 10^-2^) | 105.400 [41.400] | 23.7 \| 187.1 | 0.012 | 99.000 [39.500] | 21.1 \| 176.9 | **0.013** | <0.001 | <0.001 |
| **MULTILAYER CONNECTOME** | | | | | | | | |
| EC (x 10^-2^) | 0.600 [0.600] | -0.5 \| 1.7 | 0.290 | 0.600 [0.600] | -0.5 \| 1.7 | 0.282 | 0.401 | 0.253 |
| Ecc (x 10^-2^) | -1.600 [0.700] | -2.9 \| -0.3 | 0.013 | -1.600 [0.600] | -2.9 \| -0.4 | **0.013** | <0.001 | <0.001 |
| **RESPONDERS vs NON-RESPONDERS** | | | | | | | | |
| **FUNCTIONAL CONNECTOME** | | | | | | | | |
| Eglob (x 10^-3^) | -2.500 [1.600] | -5.6 \| 0.6 | 0.117 | -2.500 [1.600] | -5.6 \| 0.6 | 0.119 | 0.073 | 0.007 |
| Q_yeo_ (x 10^-2^) | 1.800 [0.900] | 0.1 \| 3.5 | 0.042 | 1.900 [0.900] | 0.3 \| 3.6 | **0.025** | 0.003 | <0.001 |
| PC_yeo_ (x 10^-2^) | -1.300 [0.700] | -2.6 \| 0.00 | 0.054 | -1.400 [0.700] | -2.7 \| -0.1 | **0.031** | 0.005 | <0.001 |
| Small-worldness (x 10^-2^) | 12.600 [7.800] | -2.9 \| 28 | 0.110 | 13.800 [7.700] | -1.3 \| 29 | 0.074 | 0.031 | 0.001 |
| **STRUCTURAL CONNECTOME** | | | | | | | | |
| Eglob (x 10^-4^) | -0.200 [8.500] | -17 \| 16.5 | 0.977 | -0.000 [8.600] | -16.9 \| 16.9 | 0.997 | 0.71 | 1.00 |
| Q_yeo_ (x 10^-2^) | 0.100 [0.300] | -0.5 \| 0.8 | 0.707 | 0.100 [0.300] | -0.5 \| 0.8 | 0.712 | 0.813 | 0.98 |
| PC_yeo_ (x 10^-2^) | 0.100 [0.100] | -0.1 \| 0.4 | 0.275 | 0.200 [0.100] | -0.1 \| 0.4 | 0.251 | 0.342 | 0.175 |
| Small-worldness (x 10^-2^) | 37.600 [25.700] | -13.1 \| 88.2 | 0.145 | 36.800 [24.500] | -11.5 \| 85.1 | 0.135 | 0.057 | 0.01 |
| **MULTILAYER CONNECTOME** | | | | | | | | |
| EC (x 10^-2^) | 0.600 [0.400] | -0.1 \| 1.3 | 0.104 | 0.600 [0.400] | -0.1 \| 1.3 | 0.115 | 0.083 | 0.007 |
| Ecc (x 10^-2^) | -0.400 [0.400] | -1.2 \| 0.5 | 0.378 | -0.300 [0.400] | -1.1 \| 0.5 | 0.432 | 0.427 | 0.668 |
| *Corrected for age and sex. Abbreviations: LOSO = leave-one-sample-out – either one of the trials or one of the treatments. Eglob = global efficiency, Q = modularity quality index, PC = participation coefficient, EC = eigenvector centrality, Ecc = eccentricity. | | | | | | | | |

| **Supplementary Table 5 – pre-treatment global connectome measures and treatment response - adjusting for treatment or trial** | | | | | | |
| --- | --- | --- | --- | --- | --- | --- |
|  | **Adjusted for treatment** | | | **Adjusted for trial** | | |
|  | B [SE] | 95% CI | P | B [SE] | 95% CI | P |
| **% CHANGE** | | | | | | |
| **FUNCTIONAL CONNECTOME** | | | | | | |
| Eglob (x 10^-3^) | -4.500 [2.600] | -9.5 \| 0.4 | 0.077 | -4.500 [2.600] | -9.5 \| 0.4 | 0.077 |
| Q_yeo_ (x 10^-2^) | 2.500 [1.400] | -0.2 \| 5.2 | 0.074 | 2.300 [1.400] | -0.4 \| 5.1 | 0.101 |
| PC_yeo_ (x 10^-2^) | -2.000 [1.100] | -4.1 \| 0.1 | 0.059 | -1.900 [1.100] | -4.1 \| 0.1 | 0.073 |
| Small-worldness (x 10^-2^) | 23.700 [12.400] | -0.3 \| 48.6 | 0.058 | 25.300 [12.400] | 1.2 \| 49.5 | 0.043 |
| Q_subj_ (x 10^-2^) | 1.000 [0.800] | -0.5 \| 2.5 | 0.193 | 1.100 [0.700] | -0.4 \| 2.5 | 0.147 |
| PC_sub_j (x 10^-2^) | -0.800 [0.900] | -2.6 \| 1 | 0.399 | -0.800 [0.900] | -2.6 \| 1 | 0.363 |
| **STRUCTURAL CONNECTOME** | | | | | | |
| Eglob (x 10^-4^) | 11.000 [13.900] | -16.2 \| 38.1 | 0.433 | 11.000 [13.900] | -16.2 \| 38.1 | 0.433 |
| Q_yeo_ (x 10^-2^) | 0.300 [0.500] | -0.8 \| 1.3 | 0.613 | 0.300 [0.500] | -0.8 \| 1.3 | 0.570 |
| PC_yeo_ (x 10^-2^) | 0.100 [0.200] | -0.3 \| 0.5 | 0.565 | 0.100 [0.200] | -0.3 \| 0.5 | 0.565 |
| Small-worldness (x 10^-2^) | 99.400 [39.500] | 22.1 \| 176.5 | 0.013 | 101.000 [39.500] | 22.1 \| 175.9 | 0.011 |
| Q_subj_ (x 10^-2^) | -0.000 [0.200] | -0.4 \| 0.4 | 0.964 | -0.000 [0.200] | -0.4 \| 0.4 | 0.950 |
| PC_sub_j (x 10^-2^) | -0.200 [0.400] | -0.9 \| 0.5 | 0.496 | -0.200 [0.400] | -0.9 \| 0.5 | 0.504 |
| **MULTILAYER CONNECTOME** | | | | | | |
| EC (x 10^-2^) | 0.600 [0.600] | -0.5 \| 1.7 | 0.292 | 0.500 [0.600] | -0.6 \| 1.6 | 0.421 |
| Ecc (x 10^-2^) | -1.600 [0.600] | -2.9 \| -0.4 | 0.014 | -1.600 [0.600] | -2.9 \| -0.4 | 0.013 |
| **RESPONDERS vs NON-RESPONDERS** | | | | | | |
| **FUNCTIONAL CONNECTOME** | | | | | | |
| Eglob (x 10^-3^) | -2.500 [1.600] | -5.6 \| 0.6 | 0.118 | -2.500 [1.600] | -5.6 \| 0.6 | 0.119 |
| Q_yeo_ (x 10^-2^) | 1.900 [0.900] | 0.3 \| 3.6 | 0.025 | 1.800 [0.900] | 0.2 \| 3.6 | 0.039 |
| PC_yeo_ (x 10^-2^) | -1.400 [0.700] | -2.7 \| -0.1 | 0.031 | -1.300 [0.700] | -2.7 \| -0.1 | 0.041 |
| Small-worldness (x 10^-2^) | 13.100 [7.700] | -1.8 \| 28.2 | 0.090 | 13.800 [7.700] | -1.1 \| 28.8 | 0.074 |
| Q_subj_ (x 10^-2^) | 0.500 [0.500] | -0.4 \| 1.4 | 0.297 | 0.600 [0.500] | -0.3 \| 1.5 | 0.235 |
| PC_sub_j (x 10^-2^) | -0.300 [0.600] | -1.5 \| 0.7 | 0.543 | -0.400 [0.600] | -1.5 \| 0.7 | 0.498 |
| **STRUCTURAL CONNECTOME** | | | | | | |
| Eglob (x 10^-4^) | -0.000 [8.600] | -16.7 \| 16.6 | 0.997 | -0.000 [8.600] | -16.7 \| 16.6 | 0.997 |
| Q_yeo_ (x 10^-2^) | 0.100 [0.300] | -0.5 \| 0.8 | 0.687 | 0.200 [0.300] | -0.5 \| 0.8 | 0.609 |
| PC_yeo_ (x 10^-2^) | 0.200 [0.100] | -0.1 \| 0.4 | 0.251 | 0.200 [0.100] | -0.1 \| 0.4 | 0.251 |
| Small-worldness (x 10^-2^) | 36.800 [24.500] | -10.9 \| 84.4 | 0.135 | 38.000 [24.500] | -10.9 \| 84.4 | 0.123 |
| Q_subj_ (x 10^-2^) | -0.100 [0.100] | -0.4 \| 0.1 | 0.310 | -0.100 [0.100] | -0.4 \| 0.1 | 0.331 |
| PC_sub_j (x 10^-2^) | 0.000 [0.200] | -0.4 \| 0.4 | 0.920 | 0.000 [0.200] | -0.4 \| 0.5 | 0.944 |
| **MULTILAYER CONNECTOME** | | | | | | |
| EC (x 10^-2^) | 0.600 [0.400] | -0.1 \| 1.3 | 0.118 | 0.500 [0.400] | -0.2 \| 1.2 | 0.182 |
| Ecc (x 10^-2^) | -0.300 [0.400] | -1.1 \| 0.5 | 0.484 | -0.300 [0.400] | -1.1 \| 0.5 | 0.432 |

| **Supplementary Table 6 – pre-treatment global connectome measures and treatment response using individualized community parcellations** | | | | | | | | |
| --- | --- | --- | --- | --- | --- | --- | --- | --- |
|  | **crude model** | | | **adjusted model*** | | | **LOSO** | |
|  | B [SE] | 95% CI | P | B [SE] | 95% CI | P | Trial  (harm.P) | Treatment  (harm. P) |
| **% CHANGE** | | | | | | |  |  |
| **FUNCTIONAL CONNECTOME** | | | | | | | | |
| Q_subj_ (x 10^-2^) | 1.000 [0.800] | -0.5 \| 2.5 | 0.192 | 1.100 [0.800] | -0.4 \| 2.6 | 0.155 | 0.111 | 0.03 |
| PC_subj_ (x 10^-2^) | -0.700 [0.900] | -2.6 \| 1.1 | 0.421 | -0.800 [0.900] | -2.6 \| 1 | 0.369 | 0.528 | 0.552 |
| **STRUCTURAL CONNECTOME** | | | | | | | | |
| Q_subj_ (x 10^-2^) | -0.000 [0.200] | -0.4 \| 0.4 | 0.975 | -0.000 [0.200] | -0.4 \| 0.4 | 0.877 | 0.995 | 0.995 |
| PC_subj_ (x 10^-2^) | -0.200 [0.400] | -0.9 \| 0.5 | 0.598 | -0.200 [0.400] | -0.9 \| 0.5 | 0.617 | 0.888 | 0.888 |
| **RESPONDERS vs NON-RESPONDERS** | | | | | | | | |
| **FUNCTIONAL CONNECTOME** | | | | | | | | |
| Q_subj_ (x 10^-2^) | 0.500 [0.500] | -0.4 \| 1.4 | 0.305 | 0.600 [0.500] | -0.4 \| 1.5 | 0.233 | 0.255 | 0.15 |
| PC_subj_ (x 10^-2^) | -0.300 [0.600] | -1.5 \| 0.8 | 0.577 | -0.400 [0.600] | -1.5 \| 0.7 | 0.502 | 0.763 | 0.862 |
| **STRUCTURAL CONNECTOME** | | | | | | | | |
| Q_subj_ (x 10^-2^) | -0.100 [0.100] | -0.4 \| 0.1 | 0.313 | -0.100 [0.100] | -0.4 \| 0.1 | 0.310 | 0.995 | 0.995 |
| PC_subj_ (x 10^-2^) | 0.100 [0.200] | -0.4 \| 0.5 | 0.809 | 0.000 [0.200] | -0.4 \| 0.5 | 0.851 | 0.888 | 0.888 |
| *Corrected for age and sex. Abbreviations: LOSO = leave-one-sample-out – either one of the trials or one of the treatments. Qsubj = modularity quality index using subject-specific communities , PCsubj = participation coefficient. Additionally adjusting for trial or treatment using a random intercept had little effect on these results (data not shown) | | | | | | | | |

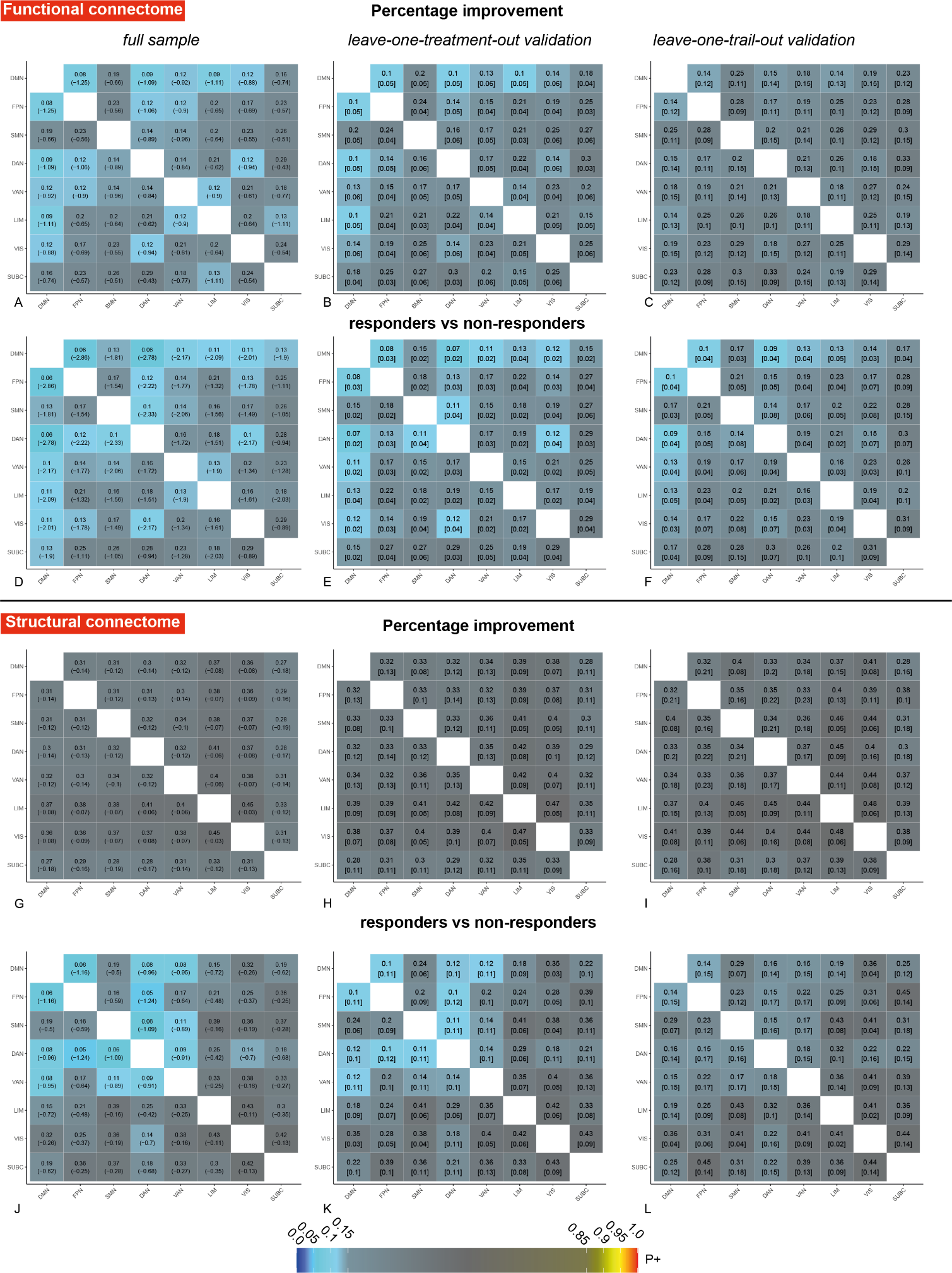

**Supplementary Figure 4- Associations between treatment efficacy and pre-treatment system-system connectivity of the functional and structural connectome.** Matrix-based Bayesian analyses showing associations between percentage improvement and functional (a-c) or structural (g-i) connectivity of 8 systems or differences between responders and non-responders in functional (d-f) or structural (j-l) connectivity. See the legend of supplementary Figure 2 for an explanation of the colors and values in the squares. Results indicated moderate evidence for negative associations between percentage improvement and functional – not structural – connectivity of the DMN with the FPN, DAN and limbic network (all P+ <0.1). Similar results were shown for responders versus non-responders. There was also credible evidence for lower structural connectivity in responders compared with non-responders for the DMN-FPN, DAN-VAN, DAN-FPN and VAN-SMN. Leave-one-treatment and leave-one-trial out validation showed that the results on DMN-FPN functional and structural connectivity were most robust. Abbreviations: DMN = default mode network, FPN = frontoparietal network, SMN = somatomotor network, DAN = dorsal attention network, VAN = ventral attention network, LIM = limbic network, VIS = visual network. SUBC = subcortical structures

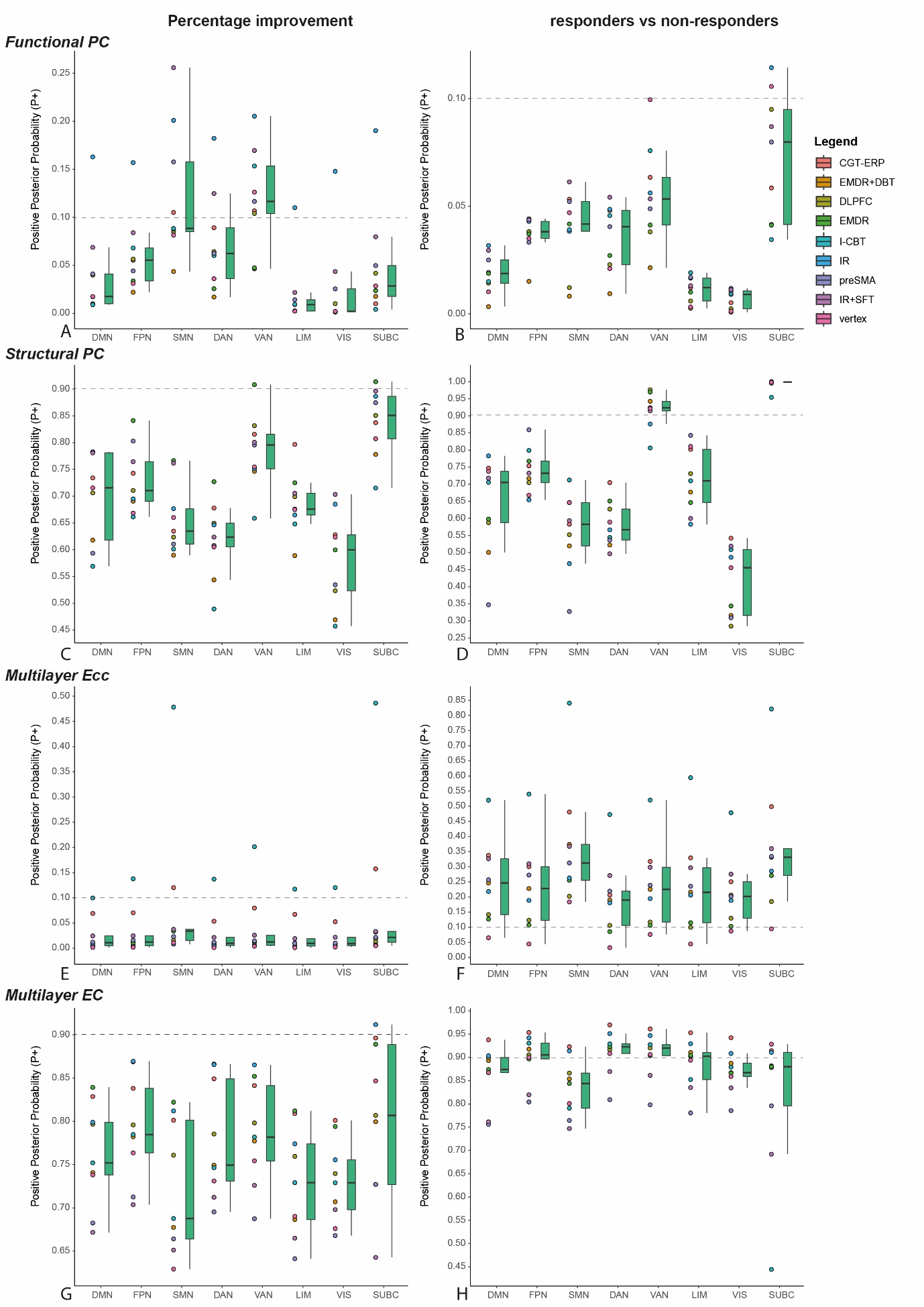

**Supplementary figure 5 – boxplots of the positive posterior probability (P+) values across leave-one-treatment-out validations.** Boxplots show the median and interquartile range of the P+ values across folds for the associations of percentage improvement (left column) with functional (a) and structural (c) participation coefficient, (e) multilayer eccentricity and (g) multilayer eigenvector centrality | (right column) P+ values for differences between responders and non-responders in functional (b) and structural (d) participation coefficient, (f) multilayer eccentricity and (h) multilayer eigenvector centrality. The dotted horizontal line indicates the P+ 0.10 or 0.90 threshold for the classification of credible evidence. Abbreviations: DMN = default mode network, FPN = frontoparietal network, SMN = somatomotor network, DAN = dorsal attention network, VAN = ventral attention network, LIM = limbic network, VIS = visual network. SUBC = subcortical structures. CBT-ERP = cognitive behavioral therapy (CBT) with exposure and response prevention (ERP) therapy, I-CBT = inference based CBT, DLPFC = repetitive Transcranial Magnetic Stimulation (rTMS) to the dorsolateral prefrontal cortex in combination with ERP, preSMA = rTMS to the pre-supplementary motor area in combination with ERP, vertex = rTMS to the vertex in combination with ERP, EMDR = eye-movement desensitization and reprocessing, EMDR+DBT = EMDR and dialectical behavioral therapy, IR = imagery rescripting therapy, IR+SFT = imagery rescripting and schema focused therapy.

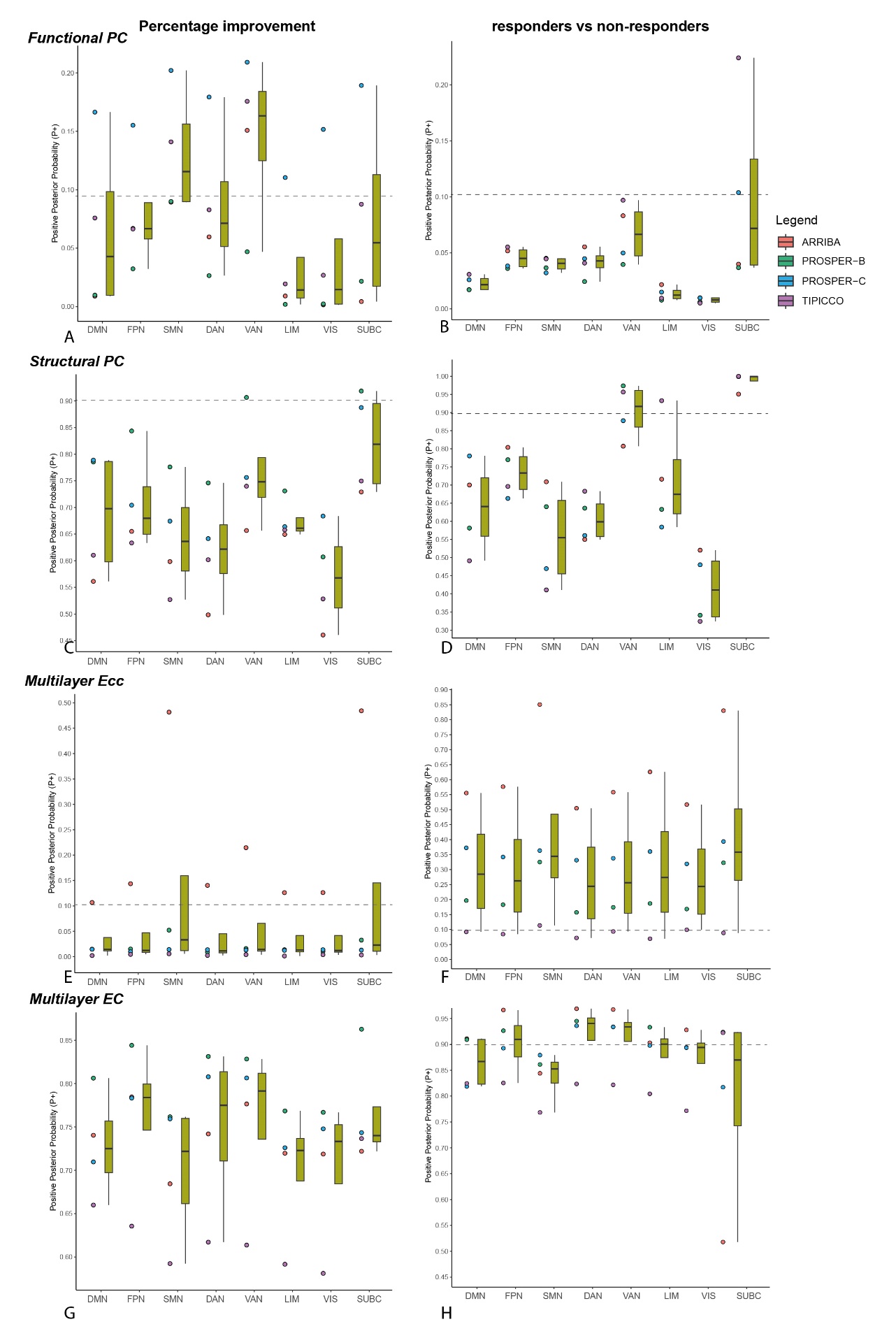

**Supplementary figure 6 – boxplots of the positive posterior probability (P+) values across leave-one-trial-out validations.** Boxplots show the median and interquartile range of the P+ values across folds for the associations of percentage improvement (left column) with functional (a) and structural (c) participation coefficient, (e) multilayer eccentricity and (g) multilayer eigenvector centrality | (right column) P+ values for differences between responders and non-responders in functional (b) and structural (d) participation coefficient, (f) multilayer eccentricity and (h) multilayer eigenvector centrality. The dotted horizontal line indicates the P+ 0.10 or 0.90 threshold for the classification of credible evidence. Abbreviations: DMN = default mode network, FPN = frontoparietal network, SMN = somatomotor network, DAN = dorsal attention network, VAN = ventral attention network, LIM = limbic network, VIS = visual network. SUBC = subcortical structures

Pre-to-post treatment changes

| **Supplementary Table 7 – pre-to-post treatment change in global connectome measures and treatment response** | | | | | | | | |
| --- | --- | --- | --- | --- | --- | --- | --- | --- |
|  | **crude model** | | | **adjusted model*** | | | **LOSO** | |
| **% CHANGE** | B [SE] | 95% CI | P | B [SE] | 95% CI | P | Trial  (harm.P) | Treatment  (harm. P) |
| **FUNCTIONAL CONNECTOME** | | | | | | | | |
| Eglob (x 10^-2^) | 6.100 [9.100] | -11.9 \| 24.1 | 0.500 | - | - | - | 0.432 | 0.766 |
| Q_yeo_ (x 10^-2^) | -5.800 [9.100] | -23.8 \| 12.2 | 0.526 | - | - | - | 0.757 | 0.887 |
| PC_yeo_ (x 10^-2^) | 4.300 [9.100] | -13.7 \| 22.3 | 0.638 | - | - | - | 0.866 | 0.982 |
| Small-worldness (x 10^-2^) | -2.900 [9.100] | -20.9 \| 15.1 | 0.750 | - | - | - | 0.879 | 0.997 |
| **STRUCTURAL CONNECTOME** | | | | | | | | |
| Eglob (x 10^-2^) | 9.200 [8.800] | -8.1 \| 26.6 | 0.295 | - | - | - | 0.364 | 0.285 |
| Q_yeo_ (x 10^-2^) | 12.100 [8.800] | -5.2 \| 29.4 | 0.170 | - | - | - | 0.165 | 0.042 |
| PC_yeo_ (x 10^-2^) | 13.700 [8.700] | -3.6 \| 31 | 0.120 | - | - | - | 0.088 | 0.009 |
| Small-worldness (x 10^-2^) | 6.900 [8.800] | -10.5 \| 24.3 | 0.434 | - | - | - | 0.65 | 0.717 |
| **MULTILAYER CONNECTOME** | | | | | | | | |
| EC (x 10^-2^) | -15.300 [9.200] | -33.5 \| 2.8 | 0.097 | - | - | - | 0.055 | 0.003 |
| Ecc (x 10^-2^) | 4.500 [9.300] | -13.9 \| 22.8 | 0.630 | - | - | - | 0.74 | 0.976 |
| **RESPONDERS vs NON-RESPONDERS** | | | | | | | | |
| **FUNCTIONAL CONNECTOME** | | | | | | | | |
| Eglob (x 10^-3^) | 0.200 [0.200] | -0.3 \| 0.7 | 0.370 | 0.200 [0.200] | -0.3 \| 0.7 | 0.369 | 0.425 | 0.489 |
| Q_yeo_ (x 10^-2^) | 0.200 [0.700] | -1.1 \| 1.5 | 0.765 | 0.200 [0.600] | -1.1 \| 1.4 | 0.763 | 0.973 | 0.998 |
| PC_yeo_ (x 10^-2^) | -0.300 [0.500] | -1.3 \| 0.8 | 0.637 | -0.300 [0.500] | -1.3 \| 0.8 | 0.635 | 0.908 | 0.967 |
| Small-worldness (x 10^-2^) | -11.000 [11.800] | -34 \| 12 | 0.353 | -11.000 [11.700] | -33.8 \| 11.8 | 0.350 | 0.518 | 0.485 |
| **STRUCTURAL CONNECTOME** | | | | | | | | |
| Eglob (x 10^-4^) | 0.000 [0.100] | -0.1 \| 0.2 | 0.631 | 0.000 [0.100] | -0.1 \| 0.2 | 0.627 | 0.836 | 0.951 |
| Q_yeo_ (x 10^-2^) | -0.100 [0.400] | -0.8 \| 0.7 | 0.872 | -0.100 [0.400] | -0.8 \| 0.7 | 0.872 | 0.734 | 0.999 |
| PC_yeo_ (x 10^-2^) | -0.100 [0.200] | -0.4 \| 0.2 | 0.491 | -0.100 [0.200] | -0.4 \| 0.2 | 0.492 | 0.74 | 0.78 |
| Small-worldness (x 10^-2^) | -61.200 [25.900] | -111.7 \| -10.7 | 0.019 | -61.200 [25.900] | -111.5 \| -10.9 | **0.019** | 0.001 | <0.001 |
| **MULTILAYER CONNECTOME** | | | | | | | | |
| EC (x 10^-2^) | -1.200 [0.600] | -2.4 \| 0 | 0.054 | -1.200 [0.600] | -2.3 \| 0 | **0.047** | 0.008 | <0.001 |
| Ecc (x 10^-2^) | -0.300 [0.600] | -1.5 \| 0.9 | 0.580 | -0.300 [0.600] | -1.5 \| 0.8 | 0.580 | 0.715 | 0.943 |
| *Corrected for age and sex. Abbreviations: LOSO = leave-one-sample-out – either one of the trials or one of the treatments. Eglob = global efficiency, Q = modularity quality index, PC = participation coefficient, EC = eigenvector centrality, Ecc = eccentricity. | | | | | | | | |

| **Supplementary Table 8 – mixed model analyses of longitudinal global connectome measures - adjusting for treatment or trial** | | | | | | |
| --- | --- | --- | --- | --- | --- | --- |
|  | **Adjusted for treatment** | | | **Adjusted for trial** | | |
|  | B [SE] | 95% CI | P | B [SE] | 95% CI | P |
| **RESPONDERS vs NON-RESPONDERS** | | | | | | |
| **FUNCTIONAL CONNECTOME** | | | | | | |
| Eglob (x 10^-3^) | 0.200 [0.200] | -0.2 \| 0.7 | 0.367 | 0.200 [0.200] | -0.2 \| 0.7 | 0.368 |
| Q_yeo_ (x 10^-2^) | 0.200 [0.600] | -1 \| 1.4 | 0.764 | 0.200 [0.600] | -1 \| 1.4 | 0.762 |
| PC_yeo_ (x 10^-2^) | -0.300 [0.500] | -1.3 \| 0.8 | 0.637 | -0.300 [0.500] | -1.3 \| 0.8 | 0.634 |
| Small-worldness (x 10^-2^) | -11.000 [11.800] | -33.5 \| 11.5 | 0.353 | -11.000 [11.800] | -33.7 \| 11.7 | 0.351 |
| Q_subj_ (x 10^-2^) | 0.100 [0.500] | -0.9 \| 1.2 | 0.807 | 0.100 [0.500] | -0.9 \| 1.2 | 0.806 |
| PC_sub_j (x 10^-2^) | -0.400 [0.700] | -1.6 \| 0.9 | 0.581 | -0.400 [0.700] | -1.6 \| 0.9 | 0.581 |
| **STRUCTURAL CONNECTOME** | | | | | | |
| Eglob (x 10^-4^) | 0.000 [0.100] | -0.1 \| 0.2 | 0.631 | 0.000 [0.100] | -0.1 \| 0.2 | 0.629 |
| Q_yeo_ (x 10^-2^) | -0.100 [0.400] | -0.8 \| 0.7 | 0.871 | -0.100 [0.400] | -0.8 \| 0.7 | 0.871 |
| PC_yeo_ (x 10^-2^) | -0.100 [0.200] | -0.4 \| 0.2 | 0.494 | -0.100 [0.200] | -0.4 \| 0.2 | 0.494 |
| Small-worldness (x 10^-2^) | -61.200 [25.700] | -110.3 \| -12.1 | **0.018** | -61.200 [25.800] | -111 \| -11.4 | **0.018** |
| Q_subj_ (x 10^-2^) | 0.100 [0.100] | -0.2 \| 0.4 | 0.617 | 0.100 [0.100] | -0.2 \| 0.4 | 0.616 |
| PC_sub_j (x 10^-2^) | 0.100 [0.300] | -0.5 \| 0.7 | 0.764 | 0.100 [0.300] | -0.5 \| 0.7 | 0.762 |
| **MULTILAYER CONNECTOME** | | | | | | |
| EC (x 10^-2^) | -1.200 [0.600] | -2.3 \| 0 | **0.048** | -1.200 [0.600] | -2.3 \| 0 | **0.047** |
| Ecc (x 10^-2^) | -0.300 [0.600] | -1.5 \| 0.8 | 0.584 | -0.300 [0.600] | -1.5 \| 0.8 | 0.582 |
| Also corrected for age and sex. Eglob = global efficiency, Q = modularity quality index, PC = participation coefficient, EC = eigenvector centrality, Ecc = eccentricity. | | | | | | |

| **Supplementary Table 9 – change in global connectome measures and treatment response using individualized community parcellations** | | | | | | | | |
| --- | --- | --- | --- | --- | --- | --- | --- | --- |
|  | **crude model** | | | **adjusted model*** | | | **LOSO** | |
|  | B [SE] | 95% CI | P | B [SE] | 95% CI | P | Trial  (harm.P) | Treatment  (harm. P) |
| **% CHANGE** | | | | | | |  |  |
| **FUNCTIONAL CONNECTOME** | | | | | | | | |
| Q_subj_ (x 10^-2^) | -6.200 [9.100] | -24.2 \| 11.8 | 0.497 | - | - | - | 0.745 | 0.822 |
| PC_subj_ (x 10^-2^) | 6.300 [9.100] | -11.7 \| 24.3 | 0.490 | - | - | - | 0.725 | 0.799 |
| **STRUCTURAL CONNECTOME** | | | | | | | | |
| Q_subj_ (x 10^-2^) | -11.500 [8.800] | -28.9 \| 5.8 | 0.190 | - | - | - | 0.179 | 0.065 |
| PC_subj_ (x 10^-2^) | 9.800 [8.800] | -7.6 \| 27.1 | 0.268 | - | - | - | 0.363 | 0.221 |
| **RESPONDERS vs NON-RESPONDERS** | | | | | | | | |
| **FUNCTIONAL CONNECTOME** | | | | | | | | |
| Q_subj_ (x 10^-2^) | 0.100 [0.500] | -0.9 \| 1.2 | 0.808 | 0.100 [0.500] | -0.9 \| 1.2 | 0.806 | 0.965 | 0.999 |
| PC_subj_ (x 10^-2^) | -0.400 [0.700] | -1.7 \| 0.9 | 0.586 | -0.400 [0.700] | -1.6 \| 0.9 | 0.583 | 0.837 | 0.946 |
| **STRUCTURAL CONNECTOME** | | | | | | | | |
| Q_subj_ (x 10^-2^) | 0.100 [0.100] | -0.2 \| 0.4 | 0.615 | 0.100 [0.100] | -0.2 \| 0.4 | 0.616 | 0.845 | 0.967 |
| PC_subj_ (x 10^-2^) | 0.100 [0.300] | -0.5 \| 0.7 | 0.762 | 0.100 [0.300] | -0.5 \| 0.7 | 0.762 | 0.841 | 0.998 |
| *Corrected for age and sex. Abbreviations: LOSO = leave-one-sample-out – either one of the trials or one of the treatments. Qsubj = modularity quality index using subject-specific communities , PCsubj = participation coefficient. | | | | | | | | |

**On the mesoscale** , there was moderate evidence for an increase in functional connectivity between the FPN and DAN (P+ = 0.93) and increase in structural connectivity between the DMN and subcortical areas (P+ = 0.91) with improvement of symptoms (Supplementary Figure 7a/g). These results were not robust against LOSO validation. Responders showed credible evidence for differences in pre-to-post treatment changes in the structural – but not functional – connectivity of several networks, with responders showing increases and non-responders decreases in connectivity (supplementary figure 7j).

There was no credible evidence for an association between percentage improvement or response status and change in participation coefficient of the functional or structural systems (supplementary Figure 8). There was, however, credible evidence for an association between percentage improvement and lower multilayer eigenvector centrality of all networks except the SMN and visual network and with higher multilayer eccentricity of the limbic network (P+ = 0.93; supplementary figure 9a,c). Non-responders compared with responders showed larger increases in multilayer eigenvector centrality of all networks except the subcortical network (supplementary figure 9d). These system-level results were however relatively sensitive to LOSO validation (supplementary figures 8-11).

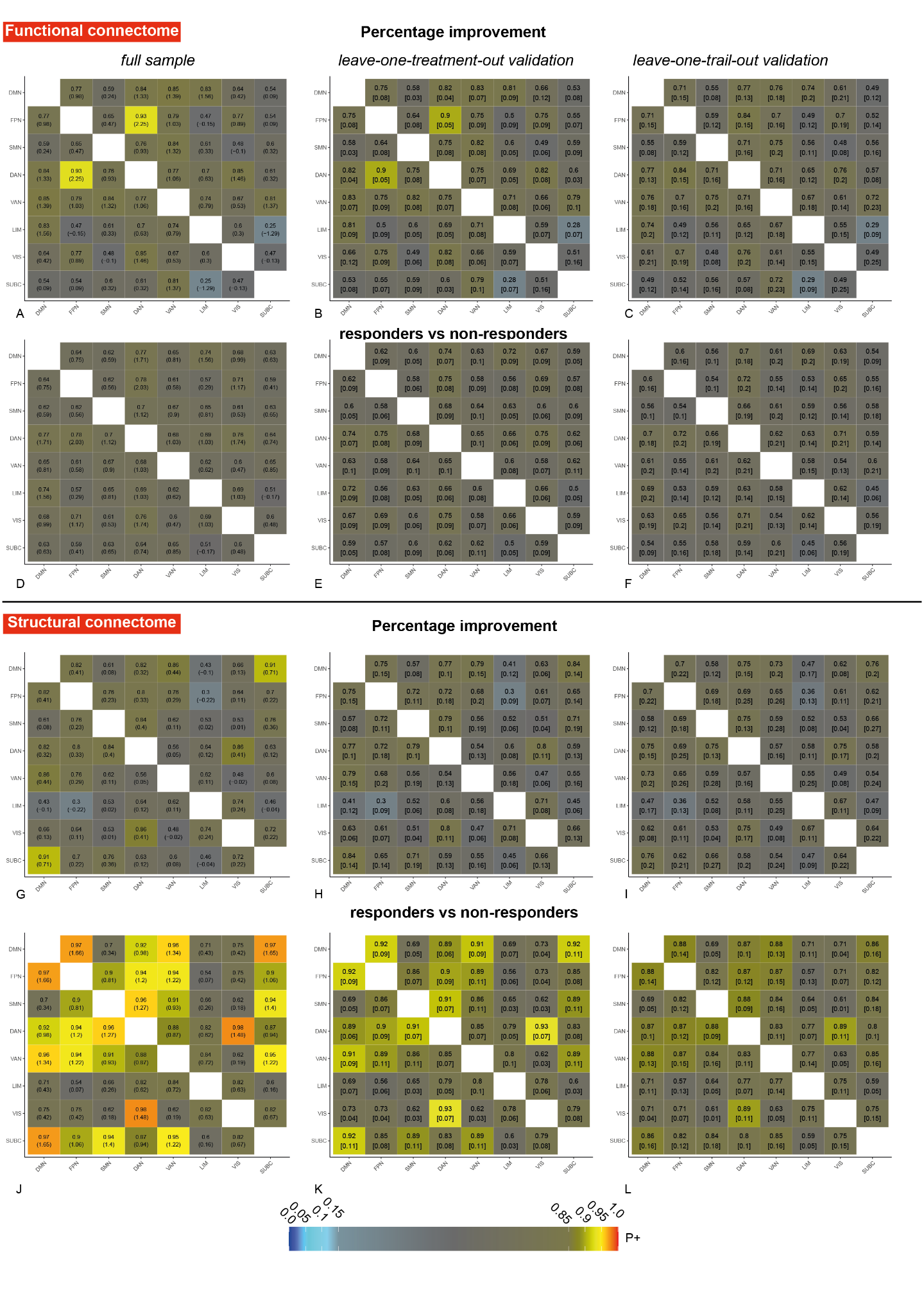

**Supplementary Figure 7- Associations between treatment efficacy and change in system-system connectivity of the functional and structural connectome from pre-to-post treatment.** Matrix-based Bayesian analyses showing associations between percentage improvement and change in functional (a-c) or structural (g-i) connectivity of 8 systems or differences between responders and non-responders in change in functional (d-f) or structural (j-l) connectivity. See the legend of supplementary Figure 2 for an explanation of the colors and values in the squares. See text above for an explanation of the results. Abbreviations: DMN = default mode network, FPN = frontoparietal network, SMN = somatomotor network, DAN = dorsal attention network, VAN = ventral attention network, LIM = limbic network, VIS = visual network. SUBC = subcortical structures

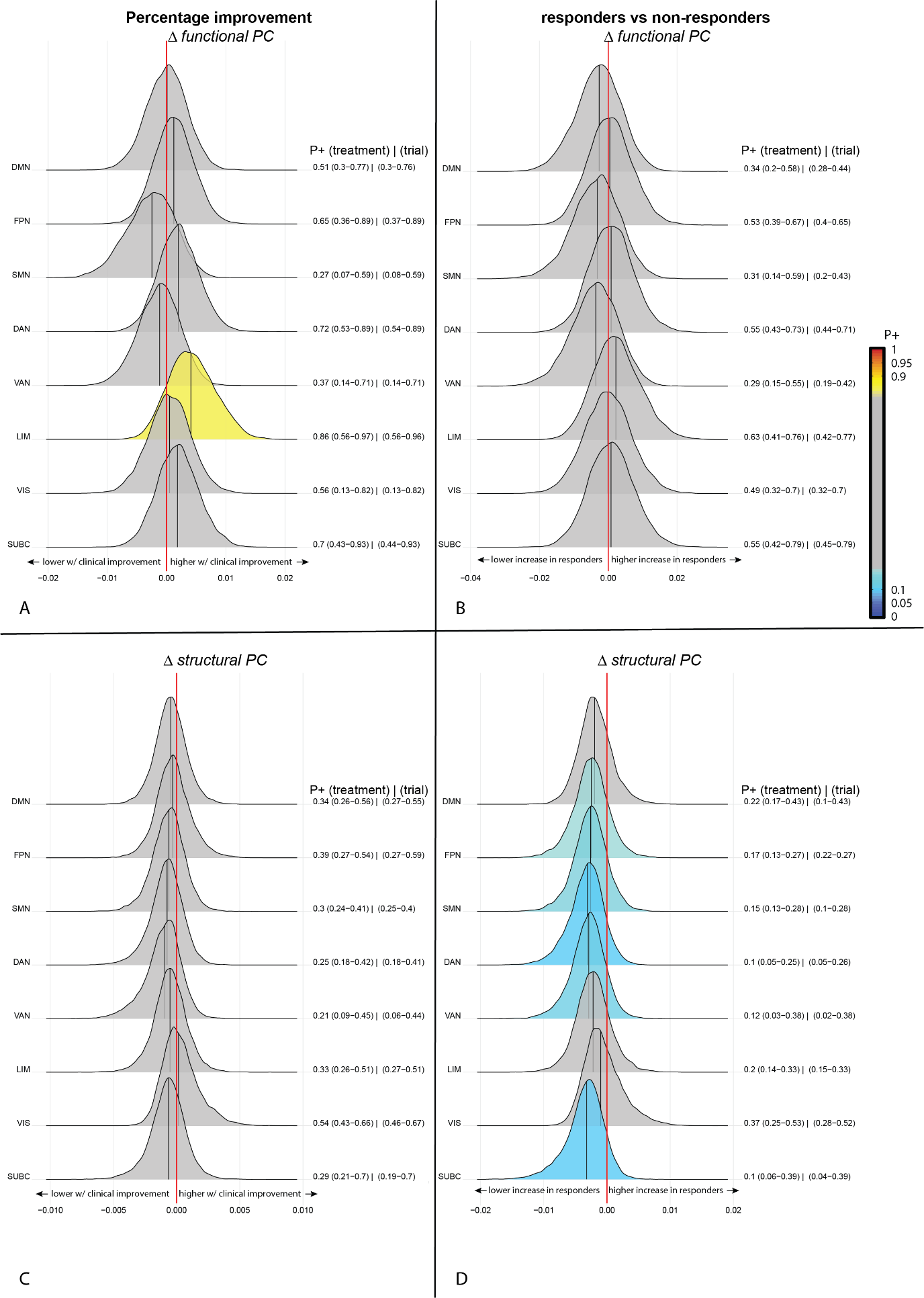

**Supplementary Figure 8 – Bayesian posterior distribution plots of the association between the pre-to-post treatment change in structural and functional participation coefficient of eight brain systems and percentage change (a,c) or response status (b,d).** The posterior distribution communicates the credibility of an effect. Positive posterior probabilities (*P+*) are shown next to each distribution and color coded. The values between brackets indicate the range of P+ values across leave-one-treatment-out, or leave-one-trial-out folds to show the robustness of the results. *P+* values ≥0.90 indicate moderate to very high credibility for a positive effect, P+ ≤0.10 indicate moderate to very high credibility for a negative effect. The meaning of the direction of effects are shown next to the red zero-effect line. See text above for an explanation of the results. abbreviations: PC = participation coefficient, DMN = default mode network, FPN = frontoparietal network, SMN = somatomotor network, DAN = dorsal attention network, VAN = ventral attention network, LIM = limbic network, VIS = visual network. SUBC = subcortical structures.

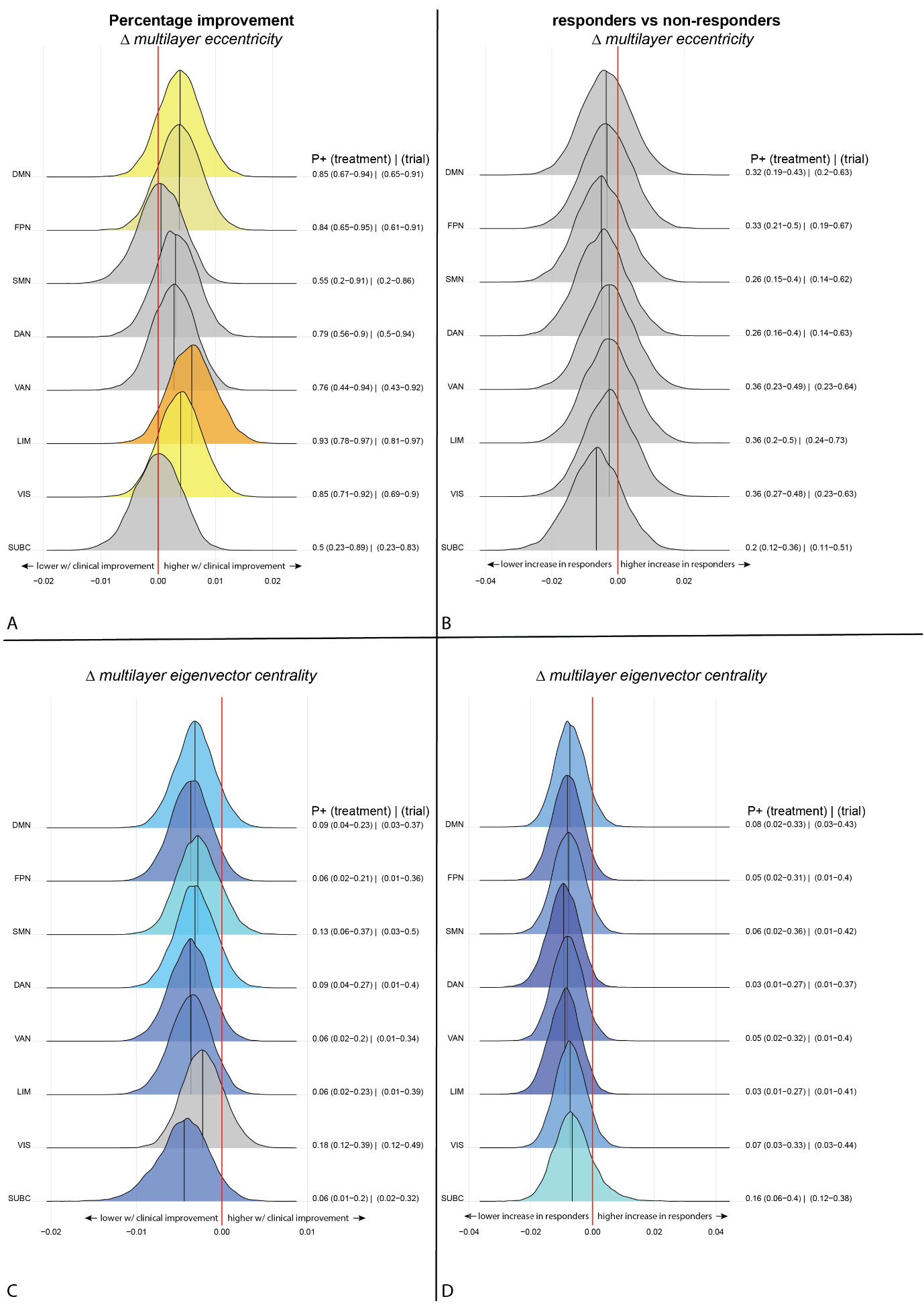

**Supplementary Figure 9 - Bayesian posterior distribution plots of the associations between the pre-to-post-treatment change in multilayer eigenvector centrality and eccentricity of eight brain systems and percentage change (a,b) or response status (c,d).** see the legend of Supplementary Figure 8 for an explanation of the posterior distribution plots and the text above for an explanation of the results. abbreviations: PC = participation coefficient, DMN = default mode network, FPN = frontoparietal network, SMN = somatomotor network, DAN = dorsal attention network, VAN = ventral attention network, LIM = limbic network, VIS = visual network. SUBC = subcortical structures.

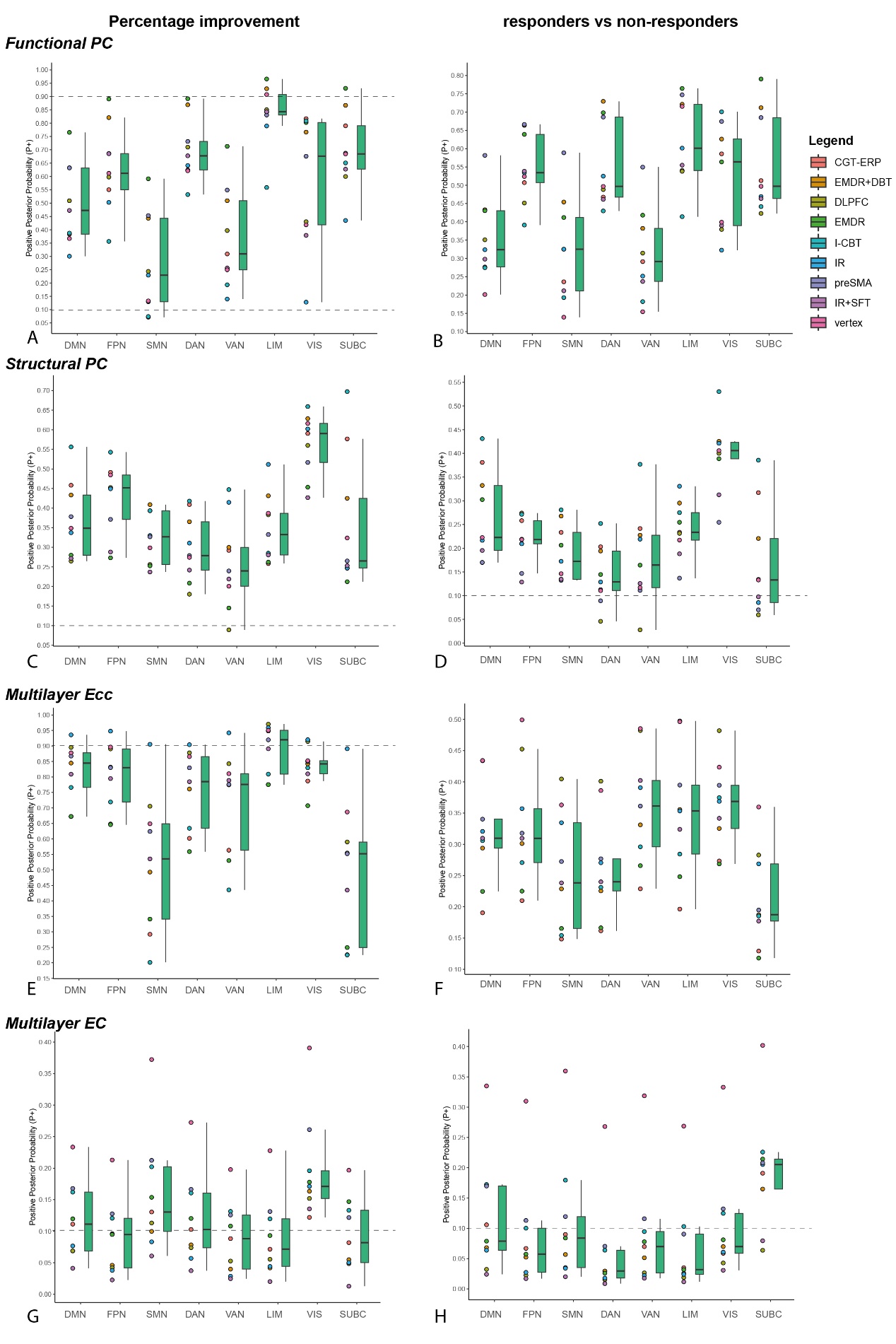

**Supplementary figure 10 – boxplots of the P+ values of associations between pre-to-post treatment changes in network topology across leave-one-treatment-out validations.** Boxplots show the median and interquartile range of the P+ values across folds for the associations of percentage improvement (left column) with pre-to-post treatment change in functional (a) and structural (c) participation coefficient, (e) multilayer eccentricity and (g) multilayer eigenvector centrality | (right column) P+ values for differences between responders and non-responders in functional (b) and structural (d) participation coefficient, (f) multilayer eccentricity and (h) multilayer eigenvector centrality. The dotted horizontal line indicates the P+ 0.10 or 0.90 threshold for the classification of credible evidence. Robustness of the results was overall low. see text above for an explanation of the results. Abbreviations: DMN = default mode network, FPN = frontoparietal network, SMN = somatomotor network, DAN = dorsal attention network, VAN = ventral attention network, LIM = limbic network, VIS = visual network. SUBC = subcortical structures. CBT-ERP = cognitive behavioral therapy (CBT) with exposure and response prevention (ERP) therapy, I-CBT = inference based CBT, DLPFC = repetitive Transcranial Magnetic Stimulation (rTMS) to the dorsolateral prefrontal cortex in combination with ERP, preSMA = rTMS to the pre-supplementary motor area in combination with ERP, vertex = rTMS to the vertex in combination with ERP, EMDR = eye-movement desensitization and reprocessing, EMDR+DBT = EMDR and dialectical behavioral therapy, IR = imagery rescripting therapy, IR+SFT = imagery rescripting and schema focused therapy.

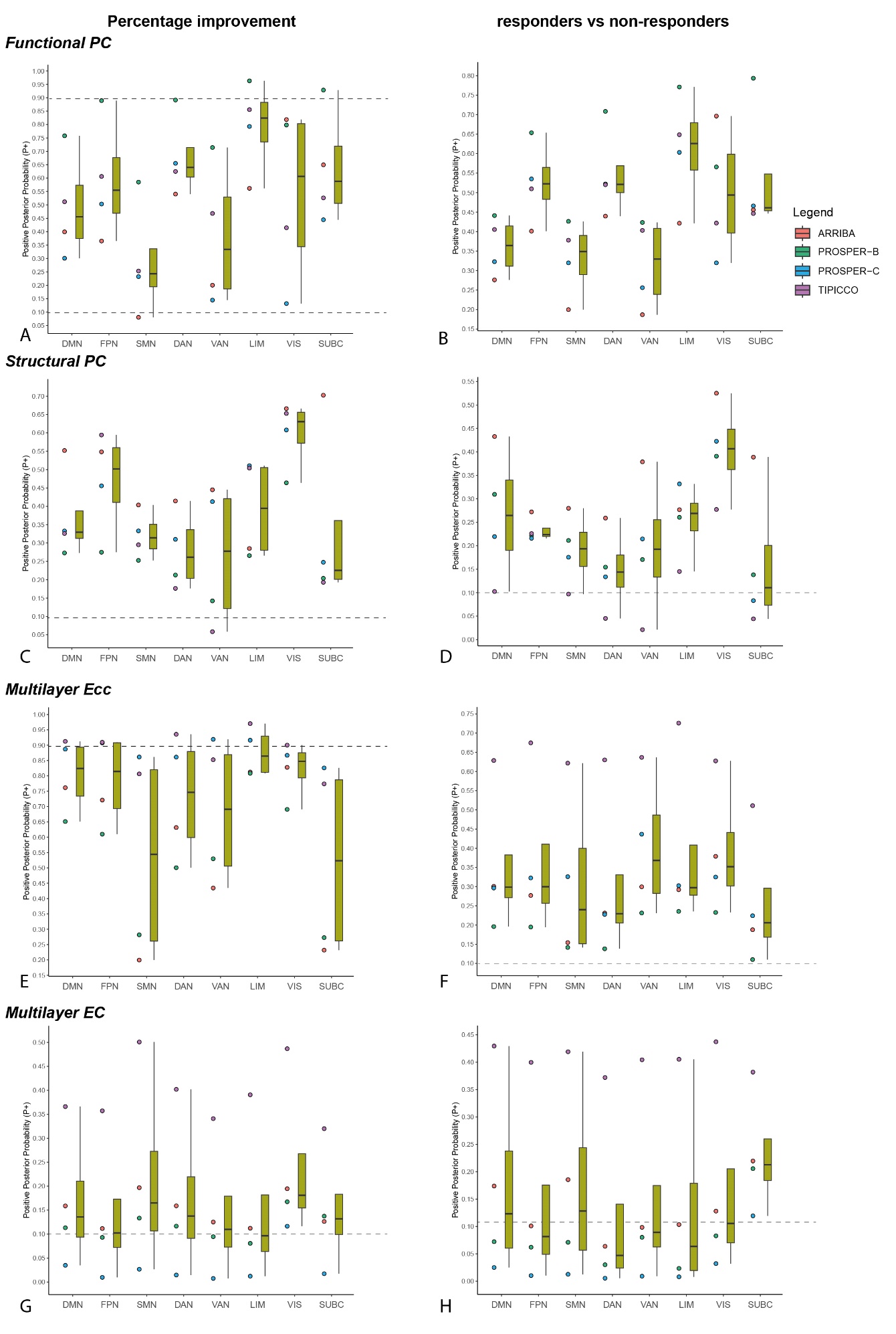

**Supplementary figure 11 – boxplots of the P+ values of associations between pre-to-post treatment changes in network topology across leave-one-trial-out validations.** Boxplots show the median and interquartile range of the P+ values across folds for the associations of percentage improvement (left column) with pre-to-post-treatment change in functional (a) and structural (c) participation coefficient, (e) multilayer eccentricity and (g) multilayer eigenvector centrality | (right column) P+ values for differences in pre-to-post-treatment change between responders and non-responders in functional (b) and structural (d) participation coefficient, (f) multilayer eccentricity and (h) multilayer eigenvector centrality. The dotted horizontal line indicates the P+ 0.10 or 0.90 threshold for the classification of credible evidence. Abbreviations: DMN = default mode network, FPN = frontoparietal network, SMN = somatomotor network, DAN = dorsal attention network, VAN = ventral attention network, LIM = limbic network, VIS = visual network. SUBC = subcortical structures

Post-hoc analyses

Adding medication status as an additional covariate to the analyses associating the pre-treatment global topological measures and treatment response did not change the results, except for the small-worldness of the functional connectome which no longer showed a significant association (see supplementary Table 10).

Bayesian analyses with leave-one-trial-out and leave-one-treatment-out validation were performed on the pre-treatment global topological measures to corroborate our NHST findings. We used the bmrs package in R (4.1.3) to perform a Bayesian regression analysis using 4 chains with 4000 iterations (1000 warmups). We choose the Student’s T distribution for the model family to account for potential outliers with weakly informative priors (normal(0,1)). These analyses showed highly credible and robust evidence for the reported associations with treatment efficacy. In addition, it showed high credibility for associations between percentage improvement and functional global efficiency (P+=0.03), average PC (P+ =0.03), modularity (P+ = 0.96) and multilayer eigenvector centrality (P+=0.90), and similar results for responder versus non-responders (supplementary Figures 12-13).

Because we included two OCD trials that used the same scale for treatment response (i.e. the YBOCS) and percentage change has been shown to be a less sensitive measure than change scores, (Vickers, 2001) we performed post-hoc analyses in the OCD samples using delta YBOCS as independent variable and included baseline YBOCS as additional covariate. On the global level, the results with the percentage change and delta-YBOCS were very similar (Supplementary Table 9). However, except for a negative association between multilayer eccentricity and change in YBOCS (B[SE]=-6.8[3.4], P=0.048), these models did not reach statistical significance. The effect sizes were similar as for the full sample suggesting that the lack of statistical significance was due to the smaller sample size.

| **Supplementary Table 10 – medication corrected mixed model analyses of global connectome measures across all trials** | | | | | | | | |
| --- | --- | --- | --- | --- | --- | --- | --- | --- |
|  | **crude model** | | | **adjusted model*** | | | **LOSO** | |
|  | B [SE] | 95% CI | P | B [SE] | 95% CI | P | Trial  (harm.P) | Treatment  (P) |
| **% CHANGE** | | | | | | |  |  |
| **FUNCTIONAL CONNECTOME** | | | | | | | | |
| Eglob (x 10^-3^) | -4.700 [2.600] | -9.8 \| 0.4 | 0.068 | -4.300 [2.600] | -9.4 \| 0.8 | 0.097 | 0.054 | 0.003 |
| Q (x 10^-2^) | 2.400 [1.400] | -0.4 \| 5.2 | 0.097 | 2.200 [1.400] | -0.5 \| 5 | 0.112 | 0.05 | 0.006 |
| PC (x 10^-2^) | -1.900 [1.100] | -4.1 \| 0.3 | 0.084 | -1.800 [1.100] | -3.9 \| 0.3 | 0.099 | 0.044 | 0.003 |
| Small-worldness (x 10^-2^) | 24.400 [12.600] | -0.5 \| 49.4 | 0.055 | 20.100 [12.300] | -4.1 \| 44.3 | 0.103 | 0.06 | 0.005 |
| **STRUCTURAL CONNECTOME** | | | | | | | | |
| Eglob (x 10^-4^) | 11.000 [13.900] | -16.3 \| 38.4 | 0.428 | 7.900 [14.100] | -19.9 \| 35.7 | 0.574 | 0.601 | 0.921 |
| Q (x 10^-2^) | 0.300 [0.500] | -0.8 \| 1.4 | 0.601 | 0.400 [0.500] | -0.7 \| 1.4 | 0.508 | 0.624 | 0.713 |
| PC (x 10^-2^) | 0.100 [0.200] | -0.3 \| 0.5 | 0.632 | 0.100 [0.200] | -0.4 \| 0.5 | 0.741 | 0.852 | 0.995 |
| Small-worldness (x 10^-2^) | 105.400 [41.400] | 23.7 \| 187.1 | 0.012 | 113.300 [38.900] | 36.5 \| 190.1 | **0.004** | <0.001 | <0.001 |
| **MULTILAYER CONNECTOME** | | | | | | | | |
| EC (x 10^-2^) | 0.600 [0.600] | -0.5 \| 1.7 | 0.290 | 0.700 [0.600] | -0.5 \| 1.8 | 0.235 | 0.317 | 0.138 |
| Ecc (x 10^-2^) | -1.600 [0.700] | -2.9 \| -0.3 | 0.013 | -1.500 [0.700] | -2.8 \| -0.2 | **0.023** | 0.001 | <0.001 |
| **RESPONDERS vs NON-RESPONDERS** | | | | | | | | |
| **FUNCTIONAL CONNECTOME** | | | | | | | | |
| Eglob (x 10^-3^) | -2.500 [1.600] | -5.6 \| 0.6 | 0.117 | -2.400 [1.600] | -5.6 \| 0.7 | 0.128 | 0.082 | 0.01 |
| Q (x 10^-2^) | 1.800 [0.900] | 0.1 \| 3.5 | 0.042 | 1.800 [0.800] | 0.1 \| 3.5 | **0.037** | 0.007 | <0.001 |
| PC (x 10^-2^) | -1.300 [0.700] | -2.6 \| 0 | 0.054 | -1.300 [0.700] | -2.6 \| 0 | **0.048** | 0.013 | <0.001 |
| Small-worldness (x 10^-2^) | 12.600 [7.800] | -2.9 \| 28 | 0.110 | 12.300 [7.500] | -2.6 \| 27.1 | 0.104 | 0.062 | 0.005 |
| **STRUCTURAL CONNECTOME** | | | | | | | | |
| Eglob (x 10^-4^) | -0.200 [8.500] | -17 \| 16.5 | 0.977 | -0.300 [8.600] | -17.2 \| 16.7 | 0.976 | 0.704 | 1 |
| Q (x 10^-2^) | 0.100 [0.300] | -0.5 \| 0.8 | 0.707 | 0.200 [0.300] | -0.5 \| 0.8 | 0.613 | 0.745 | 0.941 |
| PC (x 10^-2^) | 0.100 [0.100] | -0.1 \| 0.4 | 0.275 | 0.100 [0.100] | -0.1 \| 0.4 | 0.322 | 0.449 | 0.371 |
| Small-worldness (x 10^-2^) | 37.600 [25.700] | -13.1 \| 88.2 | 0.145 | 40.300 [24.100] | -7.2 \| 87.8 | 0.096 | 0.028 | 0.002 |
| **MULTILAYER CONNECTOME** | | | | | | | | |
| EC (x 10^-2^) | 0.600 [0.400] | -0.1 \| 1.3 | 0.104 | 0.600 [0.400] | -0.1 \| 1.3 | 0.093 | 0.056 | 0.002 |
| Ecc (x 10^-2^) | -0.400 [0.400] | -1.2 \| 0.5 | 0.378 | -0.300 [0.400] | -1.1 \| 0.5 | 0.476 | 0.464 | 0.766 |
| *Corrected for age, sex and medication status. Abbreviations: LOSO = leave-one-sample-out – either one of the trials or one of the treatments. Eglob = global efficiency, Q = modularity quality index, PC = participation coefficient, EC = eigenvector centrality, Ecc = eccentricity. Additionally adjusting for trial or treatment using a random intercept had little effect on these results. | | | | | | | | |

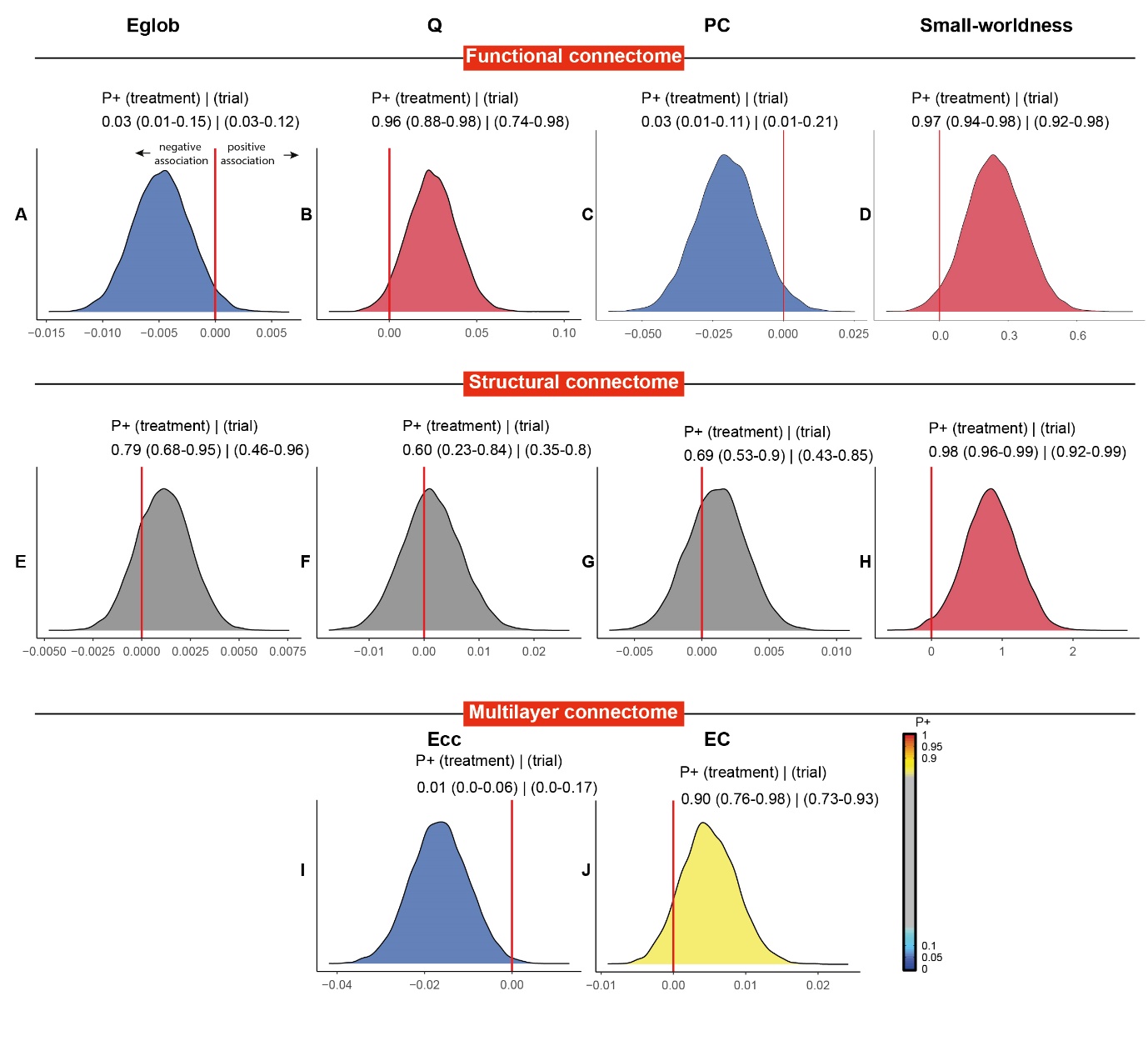

**Supplementary Figure 12 – Bayesian posterior distribution plots of the associations between percentage improvement and pre-treatment global functional structural or multilayer network measures.** see supplementary figure 3 for an explanation of the posterior distribution plots. An explanation of the direction of effect is shown in (a). Results show strongly to very strongly credible evidence for an association between percentage improvement and (a-d) functional global topology and (i) multilayer eccentricity. There was also very strongly credible evidence for an association between percentage improvement and (h) small-worldness of the structural connectome and moderate evidence for an association with multilayer eigenvector centrality. Abbreviations: Eglob = global efficiency, Q = modularity quality index, PC = participation coefficient, EC = eigenvector centrality, Ecc = eccentricity.

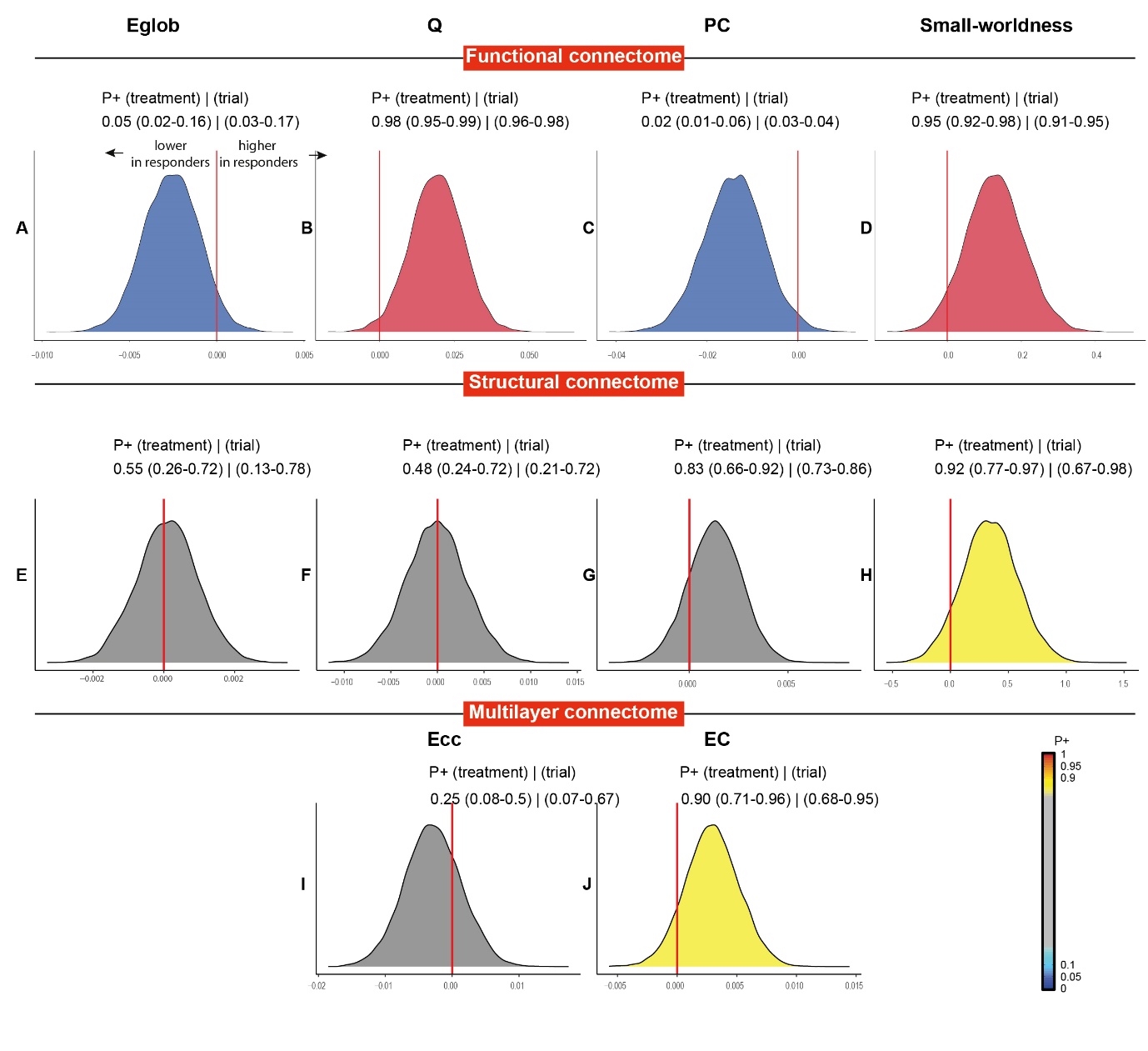

**Supplementary Figure 13 – Bayesian posterior distribution plots of the difference between responders and non-responders on pre-treatment global functional structural or multilayer network measures.** see supplementary figure 3 for an explanation of the posterior distribution plots. An explanation of the direction of effect is shown in (a). Results how strong to very strong credible evidence for differences in functional topology between responders and non-responders (a-d), and moderately strong evidence for higher structural small-worldness and multilayer eigenvector centrality in responders. Functional topology results are most robust against leave-one-treatment-out and leave-one-trial-out validation (as evidences by the P+ ranges in brackets). Abbreviations: Eglob = global efficiency, Q = modularity quality index, PC = participation coefficient, EC = eigenvector centrality, Ecc = eccentricity.

| **Supplementary Table 11 – pre-treatment global connectome measures and treatment response in OCD trials** | | | | | | |
| --- | --- | --- | --- | --- | --- | --- |
|  | **crude model#** | | | **adjusted model*** | | |
|  | B [SE] | 95% CI | P (unc) | B [SE] | 95% CI | P (unc) |
| **ΔYBOCS** | | | | | | |
| **FUNCTIONAL CONNECTOME** | | | | | | |
| Eglob (x 10^-3^) | -0.200 [0.100] | -0.5 \| 0.1 | 0.165 | -0.200 [0.100] | -0.5 \| 0.1 | 0.189 |
| Q_yeo_ (x 10^-2^) | 0.100 [0.100] | -0.1 \| 0.2 | 0.373 | 0.100 [0.100] | -0.1 \| 0.2 | 0.306 |
| PC_yeo_ (x 10^-2^) | -0.100 [0.100] | -0.2 \| 0.1 | 0.354 | -0.100 [0.100] | -0.2 \| 0 | 0.269 |
| Small-worldness (x 10^-2^) | 1.100 [0.700] | -0.4 \| 2.5 | 0.147 | 1.200 [0.700] | -0.2 \| 2.6 | 0.098 |
| **STRUCTURAL CONNECTOME** | | | | | | |
| Eglob (x 10^-4^) | 0.700 [0.800] | -0.9 \| 2.2 | 0.403 | 0.700 [0.800] | -0.9 \| 2.2 | 0.406 |
| Q_yeo_ (x 10^-2^) | -0.000 [0.000] | -0.1 \| 0 | 0.763 | -0.000 [0.000] | -0.1 \| 0 | 0.727 |
| PC_yeo_ (x 10^-2^) | 0.000 [0.000] | 0 \| 0 | 0.177 | 0.000 [0.000] | 0 \| 0 | 0.157 |
| Small-worldness (x 10^-2^) | 3.500 [2.100] | -0.7 \| 7.6 | 0.101 | 3.300 [2.000] | -0.7 \| 7.2 | 0.105 |
| **MULTILAYER CONNECTOME** | | | | | | |
| EC (x 10^-4^) | 2.200 [2.400] | -2.5 \| 6.9 | 0.359 | 2.300 [2.400] | -2.4 \| 6.9 | 0.343 |
| Ecc (x 10^-4^) | -6.800 [3.400] | -13.5 \| -0.1 | 0.048 | -6.800 [3.400] | -13.6 \| -0.1 | **0.048** |
| **% CHANGE** | | | | | | |
| **FUNCTIONAL CONNECTOME** | | | | | | |
| Eglob (x 10^-3^) | -5.300 [3.700] | -12.5 \| 2 | 0.154 | -5.100 [3.700] | -12.3 \| 2.2 | 0.170 |
| Q_yeo_ (x 10^-2^) | 1.200 [1.900] | -2.6 \| 5.1 | 0.518 | 1.300 [1.900] | -2.3 \| 5 | 0.469 |
| PC_yeo_ (x 10^-2^) | -1.100 [1.400] | -4 \| 1.7 | 0.434 | -1.300 [1.400] | -4 \| 1.5 | 0.368 |
| Small-worldness (x 10^-2^) | 26.000 [18.700] | -11.1 \| 63 | 0.167 | 28.000 [18.200] | -8 \| 64 | 0.126 |
| **STRUCTURAL CONNECTOME** | | | | | | |
| Eglob (x 10^-4^) | 7.800 [19.900] | -31.7 \| 47.2 | 0.697 | 7.600 [20.000] | -32.1 \| 47.2 | 0.707 |
| Q_yeo_ (x 10^-2^) | -0.200 [0.700] | -1.7 \| 1.2 | 0.748 | -0.300 [0.700] | -1.7 \| 1.2 | 0.725 |
| PC_yeo_ (x 10^-2^) | 0.400 [0.300] | -0.2 \| 1 | 0.195 | 0.400 [0.300] | -0.2 \| 1 | 0.179 |
| Small-worldness (x 10^-2^) | 88.700 [52.900] | -16 \| 193.4 | 0.096 | 84.800 [50.000] | -14.3 \| 183.8 | 0.093 |
| **MULTILAYER CONNECTOME** | | | | | | |
| EC (x 10^-4^) | 71.400 [59.100] | -45.7 \| 188.6 | 0.229 | 73.800 [59.100] | -43.3 \| 191 | 0.214 |
| Ecc (x 10^-4^) | -165.800 [85.300] | -334.8 \| 3.3 | 0.055 | -167.000 [85.500] | -336.6 \| 2.6 | 0.053 |
| **RESPONDERS vs NON-RESPONDERS** | | | | | | |
| **FUNCTIONAL CONNECTOME** | | | | | | |
| Eglob (x 10^-3^) | -2.200 [2.000] | -6.1 \| 1.8 | 0.282 | -2.100 [2.000] | -6.1 \| 1.9 | 0.296 |
| Q_yeo_ (x 10^-2^) | 1.200 [1.000] | -0.9 \| 3.3 | 0.253 | 1.300 [1.000] | -0.7 \| 3.3 | 0.189 |
| PC_yeo_ (x 10^-2^) | -1.000 [0.800] | -2.5 \| 0.6 | 0.228 | -1.100 [0.700] | -2.5 \| 0.4 | 0.164 |
| Small-worldness (x 10^-2^) | 10.400 [10.200] | -9.8 \| 30.7 | 0.310 | 11.200 [10.000] | -8.5 \| 30.9 | 0.264 |
| **STRUCTURAL CONNECTOME** | | | | | | |
| Eglob (x 10^-4^) | 4.200 [10.700] | -17 \| 25.5 | 0.694 | 4.700 [10.800] | -16.8 \| 26.1 | 0.668 |
| Q_yeo_ (x 10^-2^) | -0.000 [0.400] | -0.8 \| 0.8 | 0.922 | -0.000 [0.400] | -0.8 \| 0.8 | 0.956 |
| PC_yeo_ (x 10^-2^) | 0.200 [0.200] | -0.1 \| 0.5 | 0.267 | 0.200 [0.200] | -0.1 \| 0.5 | 0.287 |
| Small-worldness (x 10^-2^) | 19.300 [28.800] | -37.8 \| 76.4 | 0.505 | 22.700 [27.300] | -31.3 \| 76.7 | 0.406 |
| **MULTILAYER CONNECTOME** | | | | | | |
| EC (x 10^-4^) | 47.900 [32.400] | -16.3 \| 112.1 | 0.142 | 45.800 [32.400] | -18.5 \| 110.1 | 0.161 |
| Ecc (x 10^-4^) | -28.500 [47.600] | -122.9 \| 65.9 | 0.550 | -25.700 [47.800] | -120.5 \| 69.1 | 0.592 |
| # delta YBOCS models were additionally corrected for baseline YBOCS score. *Corrected for age and sex (and baseline YBOCS) Abbreviations: Eglob = global efficiency, Q = modularity quality index, PC = participation coefficient, EC = eigenvector centrality, Ecc = eccentricity. | | | | | | |

**REFERENCES**

Bakdash, J.Z., Marusich, L.R., 2017. Repeated Measures Correlation. Front Psychol 8, 456. https://doi.org/10.3389/fpsyg.2017.00456

Chen, G., Xiao, Y., Taylor, P.A., Rajendra, J.K., Riggins, T., Geng, F., Redcay, E., Cox, R.W., 2019. Handling multiplicity in neuroimaging through Bayesian lenses with multilevel modeling. Neuroinformatics 17, 515–545.

Dimitriadis, S.I., Salis, C., Tarnanas, I., Linden, D.E., 2017. Topological Filtering of Dynamic Functional Brain Networks Unfolds Informative Chronnectomics: A Novel Data-Driven Thresholding Scheme Based on Orthogonal Minimal Spanning Trees (OMSTs). Front. Neuroinform. 11. https://doi.org/10.3389/fninf.2017.00028

Fitzsimmons, S.M.D.D., Postma, T., van Campen, A.D., Vriend, C., Batelaan, N.M., van Oppen, P., Hoogendoorn, A.W., van der Werf, Y.D., van den Heuvel, O.A., 2024. TMS-induced plasticity improving cognitive control in OCD I: Clinical and neuroimaging outcomes from a randomised trial. Biol Psychiatry S0006-3223(24)01488–4. https://doi.org/10.1016/j.biopsych.2024.06.029

Luppi, A.I., Gellersen, H.M., Liu, Z.-Q., Peattie, A.R.D., Manktelow, A.E., Adapa, R., Owen, A.M., Naci, L., Menon, D.K., Dimitriadis, S.I., Stamatakis, E.A., 2024. Systematic evaluation of fMRI data-processing pipelines for consistent functional connectomics. Nat Commun 15, 4745. https://doi.org/10.1038/s41467-024-48781-5

McColgan, P., Blom, T., Rees, G., Seunarine, K.K., Gregory, S., Johnson, E., Durr, A., Roos, R.A., Scahill, R.I., Clark, C.A., Tabrizi, S.J., Razi, A., 2018. Stability and sensitivity of structural connectomes: effect of thresholding and filtering and demonstration in neurodegeneration. bioRxiv 416826. https://doi.org/10.1101/416826

Pouwels, P.J.W., Vriend, C., Liu, F., de Joode, N.T., Otaduy, M.C.G., Pastorello, B., Robertson, F.C., Venkatasubramanian, G., Ipser, J., Lee, S., Batistuzzo, M.C., Hoexter, M.Q., Lochner, C., Miguel, E.C., Narayanaswamy, J.C., Rao, R., Janardhan Reddy, Y.C., Shavitt, R.G., Sheshachala, K., Stein, D.J., van Balkom, A., Wall, M., Simpson, H.B., van den Heuvel, O.A., 2023. Global multi-center and multi-modal magnetic resonance imaging study of obsessive-compulsive disorder: Harmonization and monitoring of protocols in healthy volunteers and phantoms. Int J Methods Psychiatr Res 32, e1931. https://doi.org/10.1002/mpr.1931

Ran, Q., Jamoulle, T., Schaeverbeke, J., Meersmans, K., Vandenberghe, R., Dupont, P., 2020. Reproducibility of graph measures at the subject level using resting-state fMRI. Brain Behav 10, 2336–2351. https://doi.org/10.1002/brb3.1705

Satterthwaite, T.D., Elliott, M.A., Gerraty, R.T., Ruparel, K., Loughead, J., Calkins, M.E., Eickhoff, S.B., Hakonarson, H., Gur, R.C., Gur, R.E., Wolf, D.H., 2013. An improved framework for confound regression and filtering for control of motion artifact in the preprocessing of resting-state functional connectivity data. Neuroimage 64, 240–256. https://doi.org/10.1016/j.neuroimage.2012.08.052

Smith, R.E., Tournier, J.D., Calamante, F., Connelly, A., 2015. SIFT2: Enabling dense quantitative assessment of brain white matter connectivity using streamlines tractography. Neuroimage 119, 338–51. https://doi.org/10.1016/j.neuroimage.2015.06.092

Snoek, A., Beekman, A.T.F., Dekker, J., Aarts, I., van Grootheest, G., Blankers, M., Vriend, C., van den Heuvel, O., Thomaes, K., 2020. A randomized controlled trial comparing the clinical efficacy and cost-effectiveness of eye movement desensitization and reprocessing (EMDR) and integrated EMDR-Dialectical Behavioural Therapy (DBT) in the treatment of patients with post-traumatic stress disorder and comorbid (Sub)clinical borderline personality disorder: study design. BMC Psychiatry 20, 396. https://doi.org/10.1186/s12888-020-02713-x

Taylor, P.A., Reynolds, R.C., Calhoun, V., Gonzalez-Castillo, J., Handwerker, D.A., Bandettini, P.A., Mejia, A.F., Chen, G., 2023. Highlight Results, Don’t Hide Them: Enhance interpretation, reduce biases and improve reproducibility. NeuroImage 274, 120138.

van Balkom, T.D., Berendse, H.W., van der Werf, Y.D., Twisk, J.W.R., Peeters, C.F.W., Hoogendoorn, A.W., Hagen, R.H., Berk, T., van den Heuvel, O.A., Vriend, C., 2022. Effect of eight-week online cognitive training in Parkinson’s disease: A double-blind, randomized, controlled trial. Parkinsonism & Related Disorders 96, 80–87. https://doi.org/10.1016/j.parkreldis.2022.02.018

van den End, A., Beekman, A.T.F., Dekker, J., Aarts, I., Snoek, A., Blankers, M., Vriend, C., van den Heuvel, O.A., Thomaes, K., 2024. Trauma-focused and personality disorder treatment for posttraumatic stress disorder and comorbid cluster C personality disorder: a randomized clinical trial. Eur J Psychotraumatol 15, 2382652. https://doi.org/10.1080/20008066.2024.2382652

van den End, A., Dekker, J., Beekman, A.T.F., Aarts, I., Snoek, A., Blankers, M., Vriend, C., van den Heuvel, O.A., Thomaes, K., 2021. Clinical Efficacy and Cost-Effectiveness of Imagery Rescripting Only Compared to Imagery Rescripting and Schema Therapy in Adult Patients With PTSD and Comorbid Cluster C Personality Disorder: Study Design of a Randomized Controlled Trial. Front Psychiatry 12, 633614. https://doi.org/10.3389/fpsyt.2021.633614

Vickers, A.J., 2001. The use of percentage change from baseline as an outcome in a controlled trial is statistically inefficient: a simulation study. BMC Medical Research Methodology 1, 6. https://doi.org/10.1186/1471-2288-1-6

Yeo, B.T., Krienen, F.M., Sepulcre, J., Sabuncu, M.R., Lashkari, D., Hollinshead, M., Roffman, J.L., Smoller, J.W., Zollei, L., Polimeni, J.R., Fischl, B., Liu, H., Buckner, R.L., 2011. The organization of the human cerebral cortex estimated by intrinsic functional connectivity. J Neurophysiol 106, 1125–65. https://doi.org/10.1152/jn.00338.2011

**Fmriprep boilerplate**

Results included in this manuscript come from preprocessing performed using fMRIPrep 21.0.1 (Esteban, Markiewicz, et al. (2018); Esteban, Blair, et al. (2018); RRID:SCR_016216), which is based on Nipype 1.6.1 (K. Gorgolewski et al. (2011); K. J. Gorgolewski et al. (2018); RRID:SCR_002502).

*Preprocessing of B0 inhomogeneity mappings*

A total of 2 fieldmaps were found available within the input BIDS structure for this particular subject. A B0-nonuniformity map (or fieldmap) was estimated based on two (or more) echo-planar imaging (EPI) references with topup (Andersson, Skare, and Ashburner (2003); FSL 6.0.5.1:57b01774).

*Anatomical data preprocessing*

A total of 2 T1-weighted (T1w) images were found within the input BIDS dataset. All of them were corrected for intensity non-uniformity (INU) with N4BiasFieldCorrection (Tustison et al. 2010), distributed with ANTs 2.3.3 (Avants et al. 2008, RRID:SCR_004757). The T1w-reference was then skull-stripped with a Nipype implementation of the antsBrainExtraction.sh workflow (from ANTs), using OASIS30ANTs as target template. Brain tissue segmentation of cerebrospinal fluid (CSF), white-matter (WM) and gray-matter (GM) was performed on the brain-extracted T1w using fast (FSL 6.0.5.1:57b01774, RRID:SCR_002823, Zhang, Brady, and Smith 2001). A T1w-reference map was computed after registration of 2 T1w images (after INU-correction) using mri_robust_template (FreeSurfer 6.0.1, Reuter, Rosas, and Fischl 2010). Brain surfaces were reconstructed using recon-all (FreeSurfer 6.0.1, RRID:SCR_001847, Dale, Fischl, and Sereno 1999), and the brain mask estimated previously was refined with a custom variation of the method to reconcile ANTs-derived and FreeSurfer-derived segmentations of the cortical gray-matter of Mindboggle (RRID:SCR_002438, Klein et al. 2017). Volume-based spatial normalization to two standard spaces (MNI152NLin6Asym, MNI152NLin2009cAsym) was performed through nonlinear registration with antsRegistration (ANTs 2.3.3), using brain-extracted versions of both T1w reference and the T1w template. The following templates were selected for spatial normalization: FSL’s MNI ICBM 152 non-linear 6th Generation Asymmetric Average Brain Stereotaxic Registration Model [Evans et al. (2012), RRID:SCR_002823; TemplateFlow ID: MNI152NLin6Asym], ICBM 152 Nonlinear Asymmetrical template version 2009c [Fonov et al. (2009), RRID:SCR_008796; TemplateFlow ID: MNI152NLin2009cAsym].

*Functional data preprocessing*

For each of the 2 BOLD runs found per subject (across all tasks and sessions), the following preprocessing was performed. First, a reference volume and its skull-stripped version were generated using a custom methodology of fMRIPrep. Head-motion parameters with respect to the BOLD reference (transformation matrices, and six corresponding rotation and translation parameters) are estimated before any spatiotemporal filtering using mcflirt (FSL 6.0.5.1:57b01774, Jenkinson et al. 2002). The estimated fieldmap was then aligned with rigid-registration to the target EPI (echo-planar imaging) reference run. The field coefficients were mapped on to the reference EPI using the transform. BOLD runs were slice-time corrected to 1.07s (0.5 of slice acquisition range 0s-2.15s) using 3dTshift from AFNI (Cox and Hyde 1997, RRID:SCR_005927). The BOLD reference was then co-registered to the T1w reference using bbregister (FreeSurfer) which implements boundary-based registration (Greve and Fischl 2009). Co-registration was configured with six degrees of freedom. Several confounding time-series were calculated based on the preprocessed BOLD: framewise displacement (FD), DVARS and three region-wise global signals. FD was computed using two formulations following Power (absolute sum of relative motions, Power et al. (2014)) and Jenkinson (relative root mean square displacement between affines, Jenkinson et al. (2002)). FD and DVARS are calculated for each functional run, both using their implementations in Nipype (following the definitions by Power et al. 2014). The three global signals are extracted within the CSF, the WM, and the whole-brain masks. Additionally, a set of physiological regressors were extracted to allow for component-based noise correction (CompCor, Behzadi et al. 2007). Principal components are estimated after high-pass filtering the preprocessed BOLD time-series (using a discrete cosine filter with 128s cut-off) for the two CompCor variants: temporal (tCompCor) and anatomical (aCompCor). tCompCor components are then calculated from the top 2% variable voxels within the brain mask. For aCompCor, three probabilistic masks (CSF, WM and combined CSF+WM) are generated in anatomical space. The implementation differs from that of Behzadi et al. in that instead of eroding the masks by 2 pixels on BOLD space, the aCompCor masks are subtracted a mask of pixels that likely contain a volume fraction of GM. This mask is obtained by dilating a GM mask extracted from the FreeSurfer’s aseg segmentation, and it ensures components are not extracted from voxels containing a minimal fraction of GM. Finally, these masks are resampled into BOLD space and binarized by thresholding at 0.99 (as in the original implementation). Components are also calculated separately within the WM and CSF masks. For each CompCor decomposition, the k components with the largest singular values are retained, such that the retained components’ time series are sufficient to explain 50 percent of variance across the nuisance mask (CSF, WM, combined, or temporal). The remaining components are dropped from consideration. The head-motion estimates calculated in the correction step were also placed within the corresponding confounds file. The confound time series derived from head motion estimates and global signals were expanded with the inclusion of temporal derivatives and quadratic terms for each (Satterthwaite et al. 2013). Frames that exceeded a threshold of 0.5 mm FD or 1.5 standardised DVARS were annotated as motion outliers. The BOLD time-series were resampled into standard space, generating a preprocessed BOLD run in MNI152NLin6Asym space. First, a reference volume and its skull-stripped version were generated using a custom methodology of fMRIPrep. The BOLD time-series were resampled onto the following surfaces (FreeSurfer reconstruction nomenclature): fsnative, fsaverage5. Automatic removal of motion artifacts using independent component analysis (ICA-AROMA, Pruim et al. 2015) was performed on the preprocessed BOLD on MNI space time-series after removal of non-steady state volumes and spatial smoothing with an isotropic, Gaussian kernel of 6mm FWHM (full-width half-maximum). Corresponding “non-aggresively” denoised runs were produced after such smoothing. Additionally, the “aggressive” noise-regressors were collected and placed in the corresponding confounds file. All resamplings can be performed with a single interpolation step by composing all the pertinent transformations (i.e. head-motion transform matrices, susceptibility distortion correction when available, and co-registrations to anatomical and output spaces). Gridded (volumetric) resamplings were performed using antsApplyTransforms (ANTs), configured with Lanczos interpolation to minimize the smoothing effects of other kernels (Lanczos 1964). Non-gridded (surface) resamplings were performed using mri_vol2surf (FreeSurfer).

Many internal operations of fMRIPrep use Nilearn 0.8.1 (Abraham et al. 2014, RRID:SCR_001362), mostly within the functional processing workflow. For more details of the pipeline, see the section corresponding to workflows in fMRIPrep’s documentation.

*Copyright Waiver*

The above boilerplate text was automatically generated by fMRIPrep with the express intention that users should copy and paste this text into their manuscripts unchanged. It is released under the CC0 license.

*References*

Abraham, Alexandre, Fabian Pedregosa, Michael Eickenberg, Philippe Gervais, Andreas Mueller, Jean Kossaifi, Alexandre Gramfort, Bertrand Thirion, and Gael Varoquaux. 2014. “Machine Learning for Neuroimaging with Scikit-Learn.” Frontiers in Neuroinformatics 8. https://doi.org/10.3389/fninf.2014.00014.

Andersson, Jesper L. R., Stefan Skare, and John Ashburner. 2003. “How to Correct Susceptibility Distortions in Spin-Echo Echo-Planar Images: Application to Diffusion Tensor Imaging.” NeuroImage 20 (2): 870–88. https://doi.org/10.1016/S1053-8119(03)00336-7.

Avants, B. B., C. L. Epstein, M. Grossman, and J. C. Gee. 2008. “Symmetric Diffeomorphic Image Registration with Cross-Correlation: Evaluating Automated Labeling of Elderly and Neurodegenerative Brain.” Medical Image Analysis 12 (1): 26–41. https://doi.org/10.1016/j.media.2007.06.004.

Behzadi, Yashar, Khaled Restom, Joy Liau, and Thomas T. Liu. 2007. “A Component Based Noise Correction Method (CompCor) for BOLD and Perfusion Based fMRI.” NeuroImage 37 (1): 90–101. https://doi.org/10.1016/j.neuroimage.2007.04.042.

Cox, Robert W., and James S. Hyde. 1997. “Software Tools for Analysis and Visualization of fMRI Data.” NMR in Biomedicine 10 (4-5): 171–78. https://doi.org/10.1002/(SICI)1099-1492(199706/08)10:4/5<171::AID-NBM453>3.0.CO;2-L.

Dale, Anders M., Bruce Fischl, and Martin I. Sereno. 1999. “Cortical Surface-Based Analysis: I. Segmentation and Surface Reconstruction.” NeuroImage 9 (2): 179–94. https://doi.org/10.1006/nimg.1998.0395.

Esteban, Oscar, Ross Blair, Christopher J. Markiewicz, Shoshana L. Berleant, Craig Moodie, Feilong Ma, Ayse Ilkay Isik, et al. 2018. “fMRIPrep.” Software. https://doi.org/10.5281/zenodo.852659.

Esteban, Oscar, Christopher Markiewicz, Ross W Blair, Craig Moodie, Ayse Ilkay Isik, Asier Erramuzpe Aliaga, James Kent, et al. 2018. “fMRIPrep: A Robust Preprocessing Pipeline for Functional MRI.” Nature Methods. https://doi.org/10.1038/s41592-018-0235-4.

Evans, AC, AL Janke, DL Collins, and S Baillet. 2012. “Brain Templates and Atlases.” NeuroImage 62 (2): 911–22. https://doi.org/10.1016/j.neuroimage.2012.01.024.

Fonov, VS, AC Evans, RC McKinstry, CR Almli, and DL Collins. 2009. “Unbiased Nonlinear Average Age-Appropriate Brain Templates from Birth to Adulthood.” NeuroImage 47, Supplement 1: S102. https://doi.org/10.1016/S1053-8119(09)70884-5.

Gorgolewski, K., C. D. Burns, C. Madison, D. Clark, Y. O. Halchenko, M. L. Waskom, and S. Ghosh. 2011. “Nipype: A Flexible, Lightweight and Extensible Neuroimaging Data Processing Framework in Python.” Frontiers in Neuroinformatics 5: 13. https://doi.org/10.3389/fninf.2011.00013.

Gorgolewski, Krzysztof J., Oscar Esteban, Christopher J. Markiewicz, Erik Ziegler, David Gage Ellis, Michael Philipp Notter, Dorota Jarecka, et al. 2018. “Nipype.” Software. https://doi.org/10.5281/zenodo.596855.

Greve, Douglas N, and Bruce Fischl. 2009. “Accurate and Robust Brain Image Alignment Using Boundary-Based Registration.” NeuroImage 48 (1): 63–72. https://doi.org/10.1016/j.neuroimage.2009.06.060.

Jenkinson, Mark, Peter Bannister, Michael Brady, and Stephen Smith. 2002. “Improved Optimization for the Robust and Accurate Linear Registration and Motion Correction of Brain Images.” NeuroImage 17 (2): 825–41. https://doi.org/10.1006/nimg.2002.1132.

Klein, Arno, Satrajit S. Ghosh, Forrest S. Bao, Joachim Giard, Yrjö Häme, Eliezer Stavsky, Noah Lee, et al. 2017. “Mindboggling Morphometry of Human Brains.” PLOS Computational Biology 13 (2): e1005350. https://doi.org/10.1371/journal.pcbi.1005350.

Lanczos, C. 1964. “Evaluation of Noisy Data.” Journal of the Society for Industrial and Applied Mathematics Series B Numerical Analysis 1 (1): 76–85. https://doi.org/10.1137/0701007.

Power, Jonathan D., Anish Mitra, Timothy O. Laumann, Abraham Z. Snyder, Bradley L. Schlaggar, and Steven E. Petersen. 2014. “Methods to Detect, Characterize, and Remove Motion Artifact in Resting State fMRI.” NeuroImage 84 (Supplement C): 320–41. https://doi.org/10.1016/j.neuroimage.2013.08.048.

Pruim, Raimon H. R., Maarten Mennes, Daan van Rooij, Alberto Llera, Jan K. Buitelaar, and Christian F. Beckmann. 2015. “ICA-AROMA: A Robust ICA-Based Strategy for Removing Motion Artifacts from fMRI Data.” NeuroImage 112 (Supplement C): 267–77. https://doi.org/10.1016/j.neuroimage.2015.02.064.

Reuter, Martin, Herminia Diana Rosas, and Bruce Fischl. 2010. “Highly Accurate Inverse Consistent Registration: A Robust Approach.” NeuroImage 53 (4): 1181–96. https://doi.org/10.1016/j.neuroimage.2010.07.020.

Satterthwaite, Theodore D., Mark A. Elliott, Raphael T. Gerraty, Kosha Ruparel, James Loughead, Monica E. Calkins, Simon B. Eickhoff, et al. 2013. “An improved framework for confound regression and filtering for control of motion artifact in the preprocessing of resting-state functional connectivity data.” NeuroImage 64 (1): 240–56. https://doi.org/10.1016/j.neuroimage.2012.08.052.

Tustison, N. J., B. B. Avants, P. A. Cook, Y. Zheng, A. Egan, P. A. Yushkevich, and J. C. Gee. 2010. “N4itk: Improved N3 Bias Correction.” IEEE Transactions on Medical Imaging 29 (6): 1310–20. https://doi.org/10.1109/TMI.2010.2046908.

Zhang, Y., M. Brady, and S. Smith. 2001. “Segmentation of Brain MR Images Through a Hidden Markov Random Field Model and the Expectation-Maximization Algorithm.” IEEE Transactions on Medical Imaging 20 (1): 45–57. https://doi.org/10.1109/42.906424.
